# The speed of vaccination rollout and the risk of pathogen adaptation

**DOI:** 10.1101/2022.08.01.22278283

**Authors:** Sylvain Gandon, Amaury Lambert, Marina Voinson, Troy Day, Todd L. Parsons

## Abstract

Vaccination is expected to reduce disease prevalence and to halt the spread of epidemics. But pathogen adaptation may erode the efficacy of vaccination and challenge our ability to control disease spread. Here we examine the influence of the speed of vaccination rollout on the overall risk of pathogen adaptation to vaccination. We extend the framework of evolutionary epidemiology theory to account for the different steps leading to adaptation to vaccines: (1) introduction of a vaccine-escape variant by mutation from an endemic wild-type pathogen, (2) invasion of this vaccine-escape variant in spite of the risk of early extinction, (3) spread and, eventually, fixation of the vaccine-escape variant in the pathogen population. We show that the risk of pathogen adaptation is maximal for an intermediate speed of vaccination rollout. On the one hand, slower rollout decreases pathogen adaptation because selection is too weak to avoid early extinction of the new variant. On the other hand, faster rollout decreases pathogen adaptation because it reduces the influx of adaptive mutations. Hence, vaccinating faster is recommended to decrease both the number of cases and the likelihood of pathogen adaptation. We also show that pathogen adaptation is driven by its basic reproduction ratio, the efficacy of the vaccine and the effects of the vaccine-escape mutations on pathogen life-history traits. Accounting for the interplay between epidemiology, selection and genetic drift, our work clarifies the influence of vaccination policies on different steps of pathogen adaptation and allows us to anticipate the effects of public-health interventions on pathogen evolution.

**Significance statement:** Pathogen adaptation to host immunity challenges the efficacy of vaccination against infectious diseases. Are there vaccination strategies that limit the emergence and the spread of vaccine-escape variants? Our theoretical model clarifies the interplay between the timing of vaccine escape mutation events and the transient epidemiological dynamics following the start of a vaccination campaign on pathogen adaptation. We show that the risk of adaptation is maximized for intermediate vaccination coverage but can be reduced by a combination of non pharmaceutical interventions and faster vaccination rollout.

## 1 Introduction

Vaccination offers unique opportunities to protect a large fraction of the host population and thus to control spreading epidemics. In principle, comprehensive vaccination coverage can lead to pathogen eradication. In practice, however, the coverage required for eradication is often impossible to reach with imperfect vaccines [22, 45]. Moreover, pathogen adaptation may erode the efficacy of vaccination. Even if adaptation to vaccines is less common than adaptation to drugs [20, 35, 36] the spread of vaccine-escape mutations may challenge our ability to halt the spread of epidemics.

Understanding the dynamics of pathogen adaptation to vaccines is particularly relevant in the control of the ongoing SARS-CoV-2 pandemic. Yet, most theoretical studies that explore the evolution of pathogens after vaccination are based on the analysis of deterministic models and ignore the potential effects induced by the stochasticity of epidemiological dynamics. Demographic stochasticity, however, drives the intensity of genetic drift and can affect the establishment of new mutations and the long-term evolution of pathogens [55, 58, 54]. Several studies showed how the demographic stochasticity induced by finite host and pathogen population sizes alters selection on the life-history traits of pathogens [39, 32, 49]. These analytical predictions rely on the assumption that the rate of pathogen mutation is low, which allows us to decouple epidemiological and evolutionary time scales. Indeed, when the influx of new mutations is low, the new strain is always introduced after the resident pathogen population has reached its endemic equilibrium. Many pathogens, however, have relatively high mutation rates [57] and the fate of a pathogen mutant introduced away from the endemic equilibrium is likely to be affected by the dynamics of the pathogen populations. Moreover, the start of a vaccination campaign is expected to yield massive perturbations of the epidemiological dynamics and new mutations are likely to appear when the pathogen population is far from its endemic equilibrium.

The aim of the present study is to develop a versatile theoretical framework to evaluate the consequences of vaccination on the risk of pathogen adaptation to vaccination. There are six main evolutionary-epidemiological outcomes after the start of vaccination which are summarized in **Figure 1**. Some of these outcomes are more favorable than others because they do not lead to the invasion of a new variant (**Figure 1a-c**). In contrast, vaccination may result in the invasion of vaccine-escape variants (**Figure 1e-f**). In the following we use a combination of deterministic and branching process approximations to study the joint epidemiological and evolutionary dynamics of the pathogen population. This analysis reveals the importance of the speed of the vaccination rollout as well as of the life-history characteristics of the vaccine-escape variants on the probability of pathogen adaptation.

**Figure 1:**
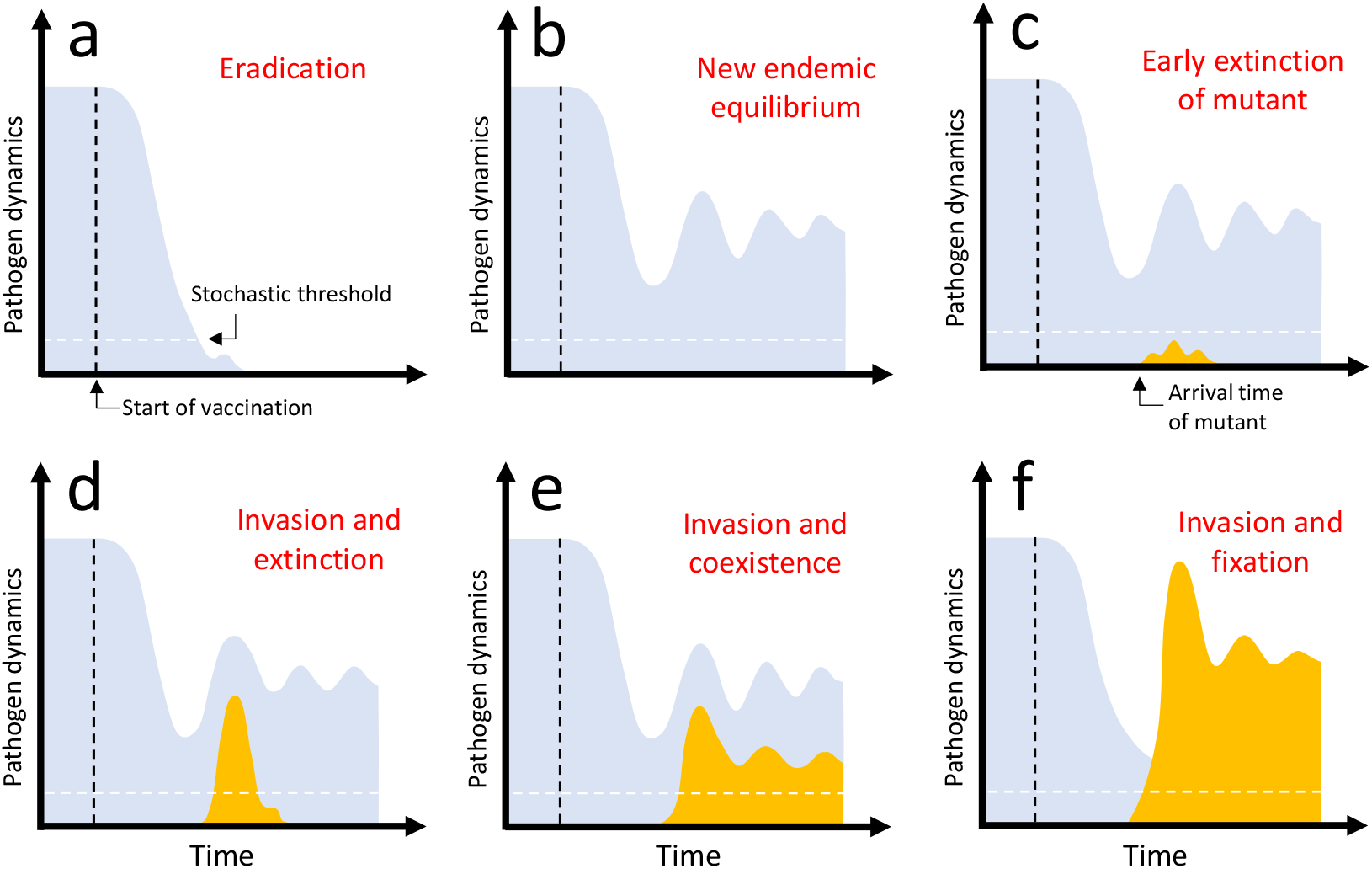
Graphical representation of the different evolutionary epidemiology outcomes after vaccination. The density of the wild-type pathogen is indicated in light blue and the dynamics of the mutant in orange. Each panel describes the temporal dynamics of the epidemics after the start of vaccination: (a) eradication of the wild-type pathogen, (b) new endemic equilibrium of the wild-type population after damped oscillations (with no introduction of the vaccine-escape mutant), (c) early extinction of the vaccine-escape mutant after its introduction by mutation, (d) invasion of the vaccine-escape mutant followed by the its extinction, (e) invasion of the vaccine-escape mutant and long-term coexistence with the wild-type in a new endemic equilibrium after damped oscillations, (f) invasion and fixation of the vaccine-escape mutant (extinction of the wild-type). The vertical dashed line (black) indicates the start of vaccination. For simplicity we consider that vaccination starts after the wild-type population has reached an endemic equilibrium. The horizontal dashed line indicates the “stochastic threshold” above which one may consider that the deterministic model provides a very good approximation of the dynamics and we can neglect the effect of demographic stochasticity. *Invasion* occurs when the vaccine-escape variant manages to go beyond the “stochastic threshold” (panels d, e and f). *Adaptation* occurs when the vaccine-escape variant is maintained in the population (panels e and f). *Fixation* occurs when the vaccine-escape variant manages to outcompete the wild-type (panel f).

## 2 Model

We use a classical SIR epidemiological model with vital dynamics (*i*.*e*., host births and deaths) [31], where hosts can be susceptible, infected or recovered [37], and are either vaccinated or unvaccinated. A host may be infected by one of two strains: a resident wild-type, or a novel mutant (we assume co-infections are not possible).

We consider a continuous-time Markov process tracking the *number* of individuals of each type of host (see Table 1 for a detailed description). Rates are interpreted as probabilities per unit time. We incorporate vital dynamics by assuming that all hosts have a base mortality rate of *δ*, while new susceptible hosts are recruited at rate *νn*. Here, *n* is a “system size”, or scaling parameter, that indicates the order of magnitude of the arena in which the epidemic occurs: the total host population varies stochastically in time, but remains of the order of *n*. We track the numbers of two classes of susceptible hosts, unvaccinated, u, or vaccinated, v, 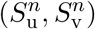, four classes of unvaccinated and vaccinated individuals, infected with the wild-type, w, 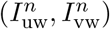 or with a mutant strain, m, 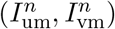, and the number recovered, *R*^*n*^. The total number susceptible is thus 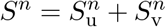, while the number of infected hosts is 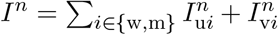. We write *H*^*n*^ for the total number of hosts:

**Table 1:**
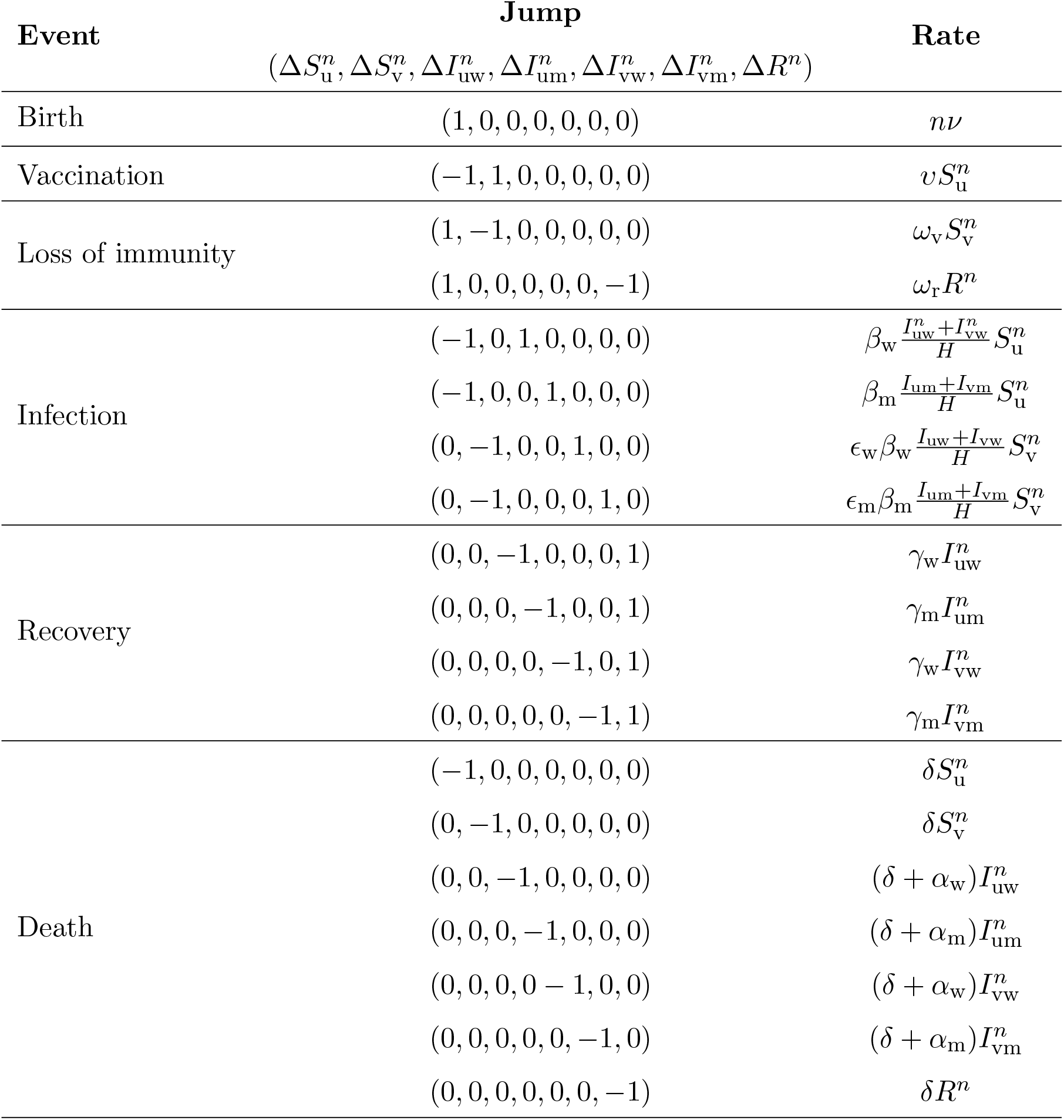
We model the epidemic via a Continuous Time Markov Chain (CTMC) with *discrete* states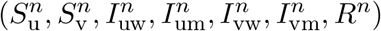. Jumps 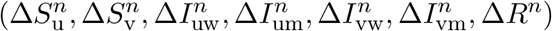 occur at state dependent rates (*i*.*e*., with probability proportional to Δ*t* in a short interval [*t, t* + Δ*t*). We implement this Markov chain using the Gillespie algorithm [25] to obtain the simulated fixation probabilities in **Figure 5** and 6.

| Event | Jump<br>$(\Delta S_u^n, \Delta S_v^n, \Delta I_{uw}^n, \Delta I_{um}^n, \Delta I_{vw}^n, \Delta I_{vm}^n, \Delta R^n)$ | Rate |
| --- | --- | --- |
| Birth | $(1, 0, 0, 0, 0, 0, 0)$ | $n\nu$ |
| Vaccination | $(-1, 1, 0, 0, 0, 0, 0)$ | $vS_u^n$ |
| Loss of immunity | $(1, -1, 0, 0, 0, 0, 0)$ | $\omega_v S_v^n$ |
| | $(1, 0, 0, 0, 0, 0, -1)$ | $\omega_r R^n$ |
| Infection | $(-1, 0, 1, 0, 0, 0, 0)$ | $\beta_w \frac{I_{uw}^n + I_{vw}^n}{H} S_u^n$ |
| | $(-1, 0, 0, 1, 0, 0, 0)$ | $\beta_m \frac{I_{um}^n + I_{vm}^n}{H} S_u^n$ |
| | $(0, -1, 0, 0, 1, 0, 0)$ | $\epsilon_w \beta_w \frac{I_{uw}^n + I_{vw}^n}{H} S_v^n$ |
| | $(0, -1, 0, 0, 0, 1, 0)$ | $\epsilon_m \beta_m \frac{I_{um}^n + I_{vm}^n}{H} S_v^n$ |
| Recovery | $(0, 0, -1, 0, 0, 0, 1)$ | $\gamma_w I_{uw}^n$ |
| | $(0, 0, 0, -1, 0, 0, 1)$ | $\gamma_m I_{um}^n$ |
| | $(0, 0, 0, 0, -1, 0, 1)$ | $\gamma_w I_{vw}^n$ |
| | $(0, 0, 0, 0, 0, -1, 1)$ | $\gamma_m I_{vm}^n$ |
| Death | $(-1, 0, 0, 0, 0, 0, 0)$ | $\delta S_u^n$ |
| | $(0, -1, 0, 0, 0, 0, 0)$ | $\delta S_v^n$ |
| | $(0, 0, -1, 0, 0, 0, 0)$ | $(\delta + \alpha_w) I_{uw}^n$ |
| | $(0, 0, 0, -1, 0, 0, 0)$ | $(\delta + \alpha_m) I_{um}^n$ |
| | $(0, 0, 0, 0, -1, 0, 0)$ | $(\delta + \alpha_w) I_{vw}^n$ |
| | $(0, 0, 0, 0, 0, -1, 0)$ | $(\delta + \alpha_m) I_{vm}^n$ |
| | $(0, 0, 0, 0, 0, 0, -1)$ | $\delta R^n$ |

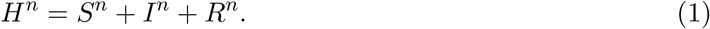

Vaccination is assumed to take place at a constant rate *υ* for all susceptible hosts. The immunity triggered by vaccination is assumed to wane at rate *ω*_v_, and natural (*i*.*e*., infection-induced) immunity is assumed to wane at rate *ω*_r_. Recovered individuals are assumed to be fully protected (no reinfections) because natural immunity is expected to be more effective than immunity triggered by vaccination (*e*.*g*., this is believed to be true for measles [8] and influenza [38, 12, 61] but not necessarily for SARS-CoV-2 [26]). We further assume that the virulence *α*_*i*_ (the mortality rate induced by the infection), the transmission *β*_*i*_ (the production rate of new infections), and the recovery *γ*_*i*_ (the rate at which the host clears the infection) are fully governed by the pathogen genotype (*i* = w or m). A fourth trait, *ϵ*_*i*_ ∈ [0, 1], governs the infectivity of pathogen genotype *i* on vaccinated hosts (infectivity of all genotypes is assumed to be equal to 1 on unvaccinated hosts). In other words, this final trait measures the ability of the pathogen to escape the immunity triggered by the vaccine. Note that these assumptions allow us to aggregate infected hosts irrespective of their vaccination status, which simplifies the analysis below. We assume frequency-dependent transmission where the number of contacts a host may have in the population is constant, but a proportion of those contacts may be infectious. Note, however, that other forms of transmission (*e*.*g*., density-dependent transmission [44]) are expected to yield qualitatively similar results. We summarize the states of the process and the jump rates at which individuals transition between states in **Table 1** and in **Figure 2**.

**Figure 2:**
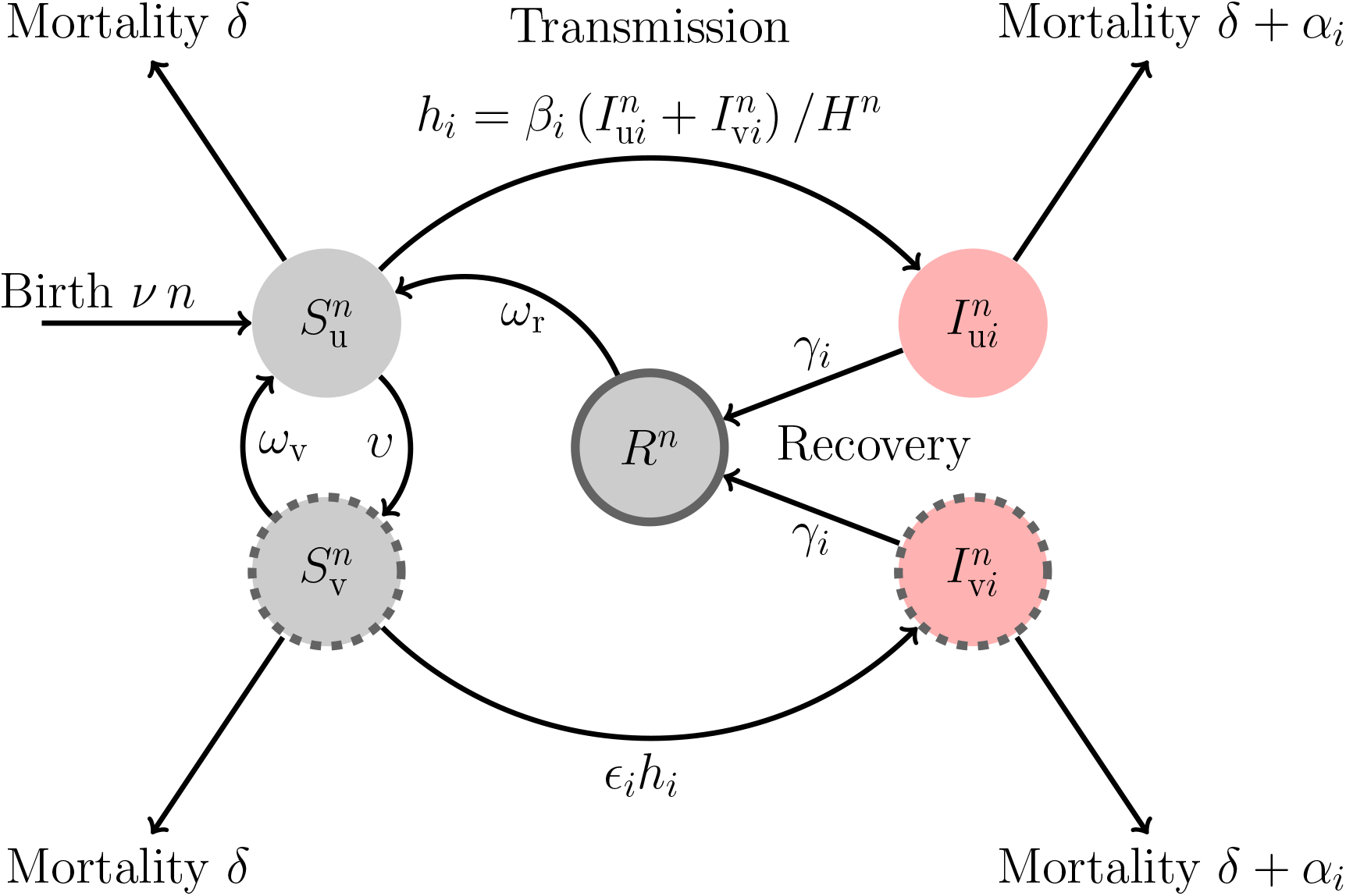
A schematic representation of the model. Naïve and uninfected hosts (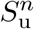 hosts) are introduced at a rate *ν* and are vaccinated at rate *υ*. Immunization induced by the vaccine wanes at rate *ω*_v_. Uninfected hosts (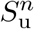 and 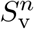) die at a rate *δ* while infected hosts (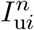 and 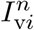) die at a rate *d*_*i*_ = *δ* + *α*_*i*_, where *i* refers to the virus genotype: the wild-type (*i* = w) or the vaccine-escape mutant (*i* = m). The rate of infection of naïve hosts by the genotype *i* is 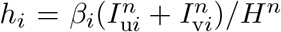, where *β*_*i*_ is the transmission rate of the genotype *i*. Vaccination reduces the force of infection and *ϵ*_*i*_ refers to the ability of the genotype *i* to escape the immunity triggered by vaccination (we assume *ϵ*_m_ *> ϵ*_w_). A host infected by pathogen genotype *i* recovers from the infection at rate *γ*_*i*_ and yields naturally immune hosts (*R*^*n*^ hosts) that cannot be reinfected by both the wild-type and the escape mutant. Natural immunity is assumed to wane at rate *ω*_r_. The total host population density is 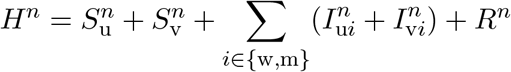.

We use this model to examine the epidemiological and evolutionary dynamics following the start of a vaccination campaign. For the sake of simplicity, we focus our analysis on scenarios where the pathogen population has reached an endemic equilibrium before the start of vaccination. This is a strong assumption, but our aim in this study is to focus on a scenario where the initial epidemiological state of the system is fixed to understand the stochastic fate of vaccine escape mutations during the transient epidemiological dynamics of the pathogen population following the start of the vaccination campaign. This is a necessary first step before studying more complex scenarios where vaccination starts before the epidemic has reached an endemic equilibrium. The default parameter values used to explore numerically the dynamics of viral adaptation are consistent with a broad range of acute infections of humans (*e*.*g*., SARS-CoV, Influenza, Measles, see **Table SI.1**). In the Discussion, we explore the robustness of our results after relaxing some of our simplifying assumptions.

## 3 Results

### 3.1 Two Approximations

Following [48], our analysis makes use of two approximations to our Markov process model. The first, deterministic approximation, uses ordinary differential equations (ODEs) and is appropriate when all types of host are abundant, but fails to correctly capture the dynamics when one or more types is rare (*e*.*g*., at the time of introduction of the mutant strain). The second uses a birth-and-death process (see *e*.*g*., [6]) to approximate rare quantities and captures stochastic phenomena, like extinction.

#### 3.1.1. Deterministic Approximation

For our first, deterministic approximation, we work with host densities defined by

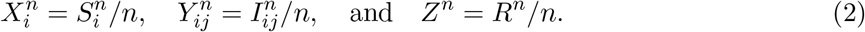

(*i* = u, v, *j* = w, m) and set

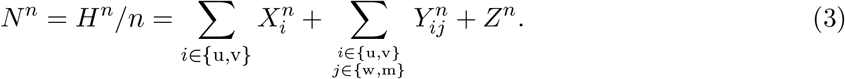

As *n* becomes large, the changes in the densities due to jumps in the Markov chain become smaller and smaller. As *n* → ∞, the 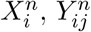, and *Z*^*n*^ approach limits *X*_*i*_, *Y*_*ij*_, and *Z*. These limits obey a system of ordinary differential equations:

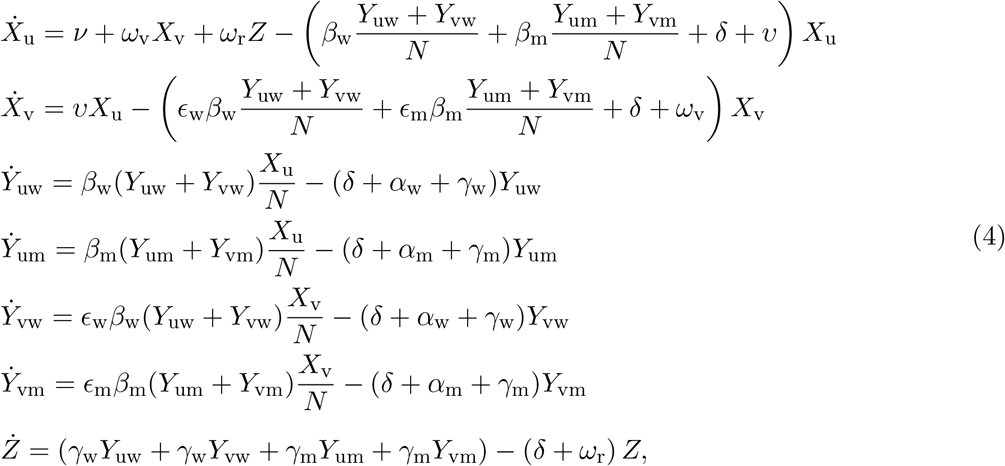

This corresponds to replacing discrete individuals by continuous densities and interpreting the rates in **Figure 2** as describing continuous flows rather than jumps (see Example B on p. 453 and Theorem 11.2.1 on p. 456 in [18] for the details and proofs of this approximation; [4] gives a readable summary with an epidemiological focus).

It is also convenient to track the dynamics of the total density of hosts infected with the same strain *i, Y*_*i*_ := *Y*_u*i*_ + *Y*_v*i*_, which yields:

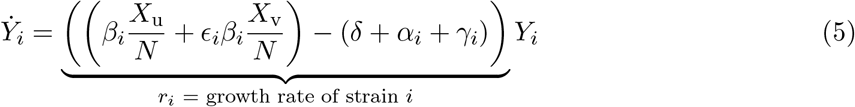

The ability of the strain *i* to grow is given by the sign of the growth rate *r*_*i*_. Note that this growth rate depends on the four different traits of the pathogen: *α*_*i*_, *β*_*i*_, *γ*_*i*_, *ϵ*_*i*_. The growth rate also depends on the densities *X*_u_(*t*) and *X*_v_(*t*), which vary with *t*, the time since the start of vaccination (*i*.*e*., vaccination starts at *t* = 0). For simplicity, we assume that at time *t* = 0, the wild-type is at its endemic equilibrium (see (SI.1) in the Supplementary Information for details), and that there are no mutants (we relax this assumption in the Supplementary Information §5).

The coefficient of selection *s*_m_(*t*) on the mutant strain relative to the wild-type is:

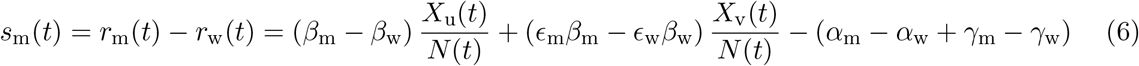

In other words, both the genetics (the phenotypic traits of strain *i*) and the environment (the epidemiological state of the host population) govern selection and strain dynamics.

##### Pathogen eradication and vaccination threshold

The ability of the strain *i* to grow can be measured by its effective per-generation reproduction ratio which is given by:

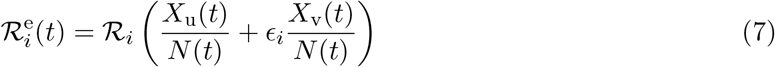

where 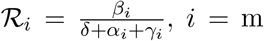, w. Hence, a reduction of the availability of susceptible hosts with vaccination may drive down the density of the wild-type pathogen when the production of new infected hosts (infection “birth”) does not compensate for the recovery and death of infected hosts (infection “death”), that is, when 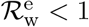. Ultimately, vaccination can even lead to the eradication of the wild-type pathogen (**Figure 1a**) either when the vaccine is sufficiently efficient (*ϵ*_w_ℛ_w_ *<* 1) or when the vaccination coverage is sufficiently high [45, 22].

Interestingly, if the aim is to eradicate an already established disease, bringing the reproduction number of the wild-type strain at the disease free equilibrium below one, (*i*.*e*., 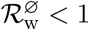 see (SI.3)), may not be sufficient to do so. Indeed, as pointed out by several earlier studies [41, 28], imperfect vaccination may yield backward bifurcation at the disease free equilibrium. In this case, the pathogen may persist even when vaccination brings 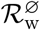 below one. Yet, the analysis of our model indicates that the condition for the emergence of backward bifurcation are very limited (see Supplementary Information §1.3) and in the following we use the condition 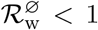 to identify the critical rate *υ*_c_ of the speed of vaccination rollout above which the wild-type pathogen can be driven to extinction (see Supplementary Information §1.3):

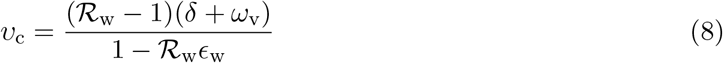

As expected, better vaccines (*i*.*e*., lower values of *ϵ*_w_ and *ω*_v_) yield lower threshold values for the speed of vaccination. Imperfect vaccines (*i*.*e*., higher values of *ϵ*_w_ and *ω*_v_), in contrast, are unlikely to allow eradication. Note that, if we wait sufficiently long, the population of the wild-type pathogen will be driven to extinction by *stochastic* fluctuations even when *υ < υ*_c_ [3, 29]. Indeed, in a finite host population, sooner or later, the pathogen population is doomed to go extinct because of demographic stochasticity, but the extinction time when *υ < υ*_c_ will usually be very long, increasing exponentially with the system size *n* [59, 5, 47]. From now on, we neglect the possibility of extinction of the wild-type due to vaccination when *υ < υ*_c_ (which is a good approximation when *n* is large).

The spread of a new pathogen variant may erode the efficacy of vaccination and, consequently, could affect the ability to control and, ultimately, to eradicate the pathogen. However, before the replacement of the wild-type by a vaccine-escape variant the pathogen population may go through three steps that may ultimately result (or not) in pathogen adaptation: (1) introduction of the vaccine-escape variant by mutation, (2) extinction (**Figure 1c**) or invasion (**Figure 1d-f**) of the vaccine-escape variant introduced by mutation, (3) fixation (**Figure 1f**) or not (**Figure 1d-e**) of the invading vaccine-escape variant. Each of these steps is very sensitive to demographic stochasticity because the number of vaccine-escape variants is very small in the early phase of its emergence. This motivates our second approximation, below.

#### 3.1.2. Birth-and-Death Process Approximation

Suppose that a mutant strain appears at time *t*_int_ ≥ 0 in a single infected host, 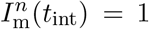, that is, with density 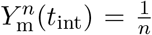. Taking *n* → ∞, we get *Y*_m_(*t*_int_) = 0. Using this as an initial condition in (4), we find that *Y*_m_(*t*) ≡ 0 for all *t* ≥ *t*_int_. This does not mean that the mutant is absent, but is simply not yet sufficiently abundant to be visible at the coarse resolution of the ODE approximation, (4). In particular, while rare, the mutant strain does not have a detectable effect on the density of susceptible hosts.

To account for the rare mutant, we use (4) to define a birth-and-death process, *Ĩ*_m_(*t*), that approximates the *number* of individuals infected with the mutant strain at times *t* ≥ *t*_int_, and allows us to estimate the probabilities of invasion (Section 3.2.2) and fixation (Section 5.1) of the mutant strain.

Each death in the birth-and-death process corresponds to the removal of a susceptible, which occurs by host death or recovery at combined rate

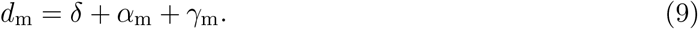

We approximate the rate of new infections,

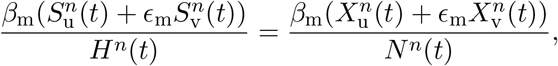

by replacing the stochastic quantities 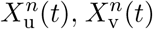 and *N*^*n*^(*t*) by their deterministic approximations *X*_u_(*t*), *X*_v_(*t*) and *N* (*t*), giving the time-dependent birth rate

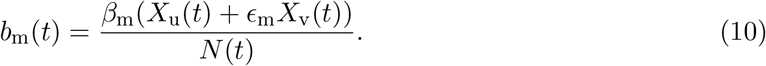

As we observed above, for the deterministic approximation, *Y*_m_(*t*_int_) = 0, and so we can compute *X*_u_(*t*), *X*_v_(*t*) and *N* (*t*) using (4) *without* the mutant strain, using initial conditions (SI.1). See [49, Supplementary Information §8.2] for a rigorous justification for this approximation.

The so-called “merciless dichotomy” [33] tells us that, started with one individual, the birth-and-death process either goes extinct, or grows indefinitely. Thus, either the mutant strain vanishes, or the number infected with the mutant strain will eventually grow to be of the order of *n* individuals, after which we can use (4) to compute the densities of both wild-type and mutant strains.

### 3.2 The Steps of Pathogen Adaptation

Using the two approximations above, we quantify the steps of pathogen evolution. First, we consider the appearance of a novel vaccine-resistant variant, which will either rapidly go extinct, or invade, causing an epidemic outbreak. Then, at the end of an epidemic, susceptible hosts are depleted, and there are few remaining infected with either wild-type and mutant strains, and both strains are at risk of extinction. If the variant outlives the wild-type, then the pathogen has adapted to the vaccine.

#### 3.2.1 Step 1: Introduction of the variant by mutation

The first step of adaptation is driven by the production of new variants of the wild-type pathogen through mutation. The degree of adaptation to unvaccinated and vaccinated hosts may vary among those variants [16]. For instance, some vaccine-escape mutations may carry no fitness costs (or may even be adaptive) in unvaccinated hosts. These variants would be expected to invade and fix because they are strongly favoured by natural selection when the proportion of vaccinated hosts builds up. They will have a strong probability to avoid the risk of early extinction irrespective of the vaccination strategy. We thus focus on variants that carry fitness costs in immunologically naïve hosts (*i*.*e*., variants *specialized* on vaccinated hosts [16]). In principle, the introduction of the vaccine-escape mutation may occur before the rollout of vaccination. The distribution of these mutations is expected to follow a stationary distribution resulting from the action of recurrent mutations and negative selection (see Supplementary Information, §5). If the fitness cost in naïve hosts is high and/or if the mutation rate is low then these pre-existing mutants are expected to be rare. In the following, we neglect the presence of pre-existing mutants and we focus on a scenario where the first vaccine-escape mutant appears after the start of vaccination (but see Supplementary Information, §5 where we discuss the effect of standing genetic variation).

At the onset of the vaccination campaign (*i*.*e*., *t* = 0) we assume that the system is at the endemic equilibrium (the equilibrium densities *X*_u_(0), *Y*_uw_(0) and *Y*_vw_(0) are given in (SI.1)). We assume that an individual host infected with the wild-type produces vaccine-escape mutants at a small, constant rate *θ*_u_*/n* if unvaccinated and *θ*_v_*/n* if vaccinated. While the rate of mutation is assumed to be constant through time, whether or not a mutant will escape extinction within a host may depend on the type of host. Indeed, a vaccine-escape mutation may have a higher probability to escape within-host extinction in vaccinated hosts. We account for this effect by making a distinction between *θ*_u_ and *θ*_v_. If vaccine-escape mutations are more likely to escape extinction in vaccinated hosts we expect *θ*_v_ *> θ*_u_. In other words, *θ*_v_*/θ*_u_ − 1 is a measure of the within-host fitness advantage of the vaccine-escape mutant in vaccinated hosts (they are assumed to have the same within-host fitness in naïve hosts). We assume that *θ*_u_ and *θ*_v_ are small enough that within-host clonal interference among vaccinated-adapted variants is negligible. The total rate of production of mutants is thus equal to

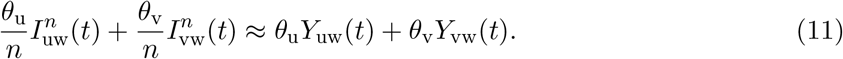

The arrival times of novel mutants are thus well approximated by a non-homogeneous Poisson process [14, p. 4] with rate

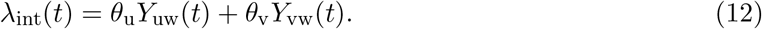

The probability that the arrival time *T*_int_ of the first vaccine-escape mutant is thus approximated by:

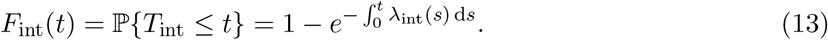

In other words, the time *T*_int_ at which the vaccine-escape variant is first introduced by mutation depends on the dynamics of the incidence of the infections by the wild-type. Plots of *F*_int_(*t*) for different values of rollout speed *υ* in **Figure 3** show that a faster rollout of vaccination delays the introduction of the vaccine-escape mutant. This effect is particularly marked when *ω*_r_ = 0 because life-long immunity is known to result in a massive transient drop of the incidence (the honey-moon period)[45, 19] which is expected to decrease the influx of new variants during this period (**Figure SI.1**). **Figure 3** also shows how higher values of *ω*_v_ can increase the influx of vaccine-escape variants. As discussed in the following section, the subsequent fate of vaccine-escape mutants depends strongly on the timing of their arrival.

**Figure 3:**
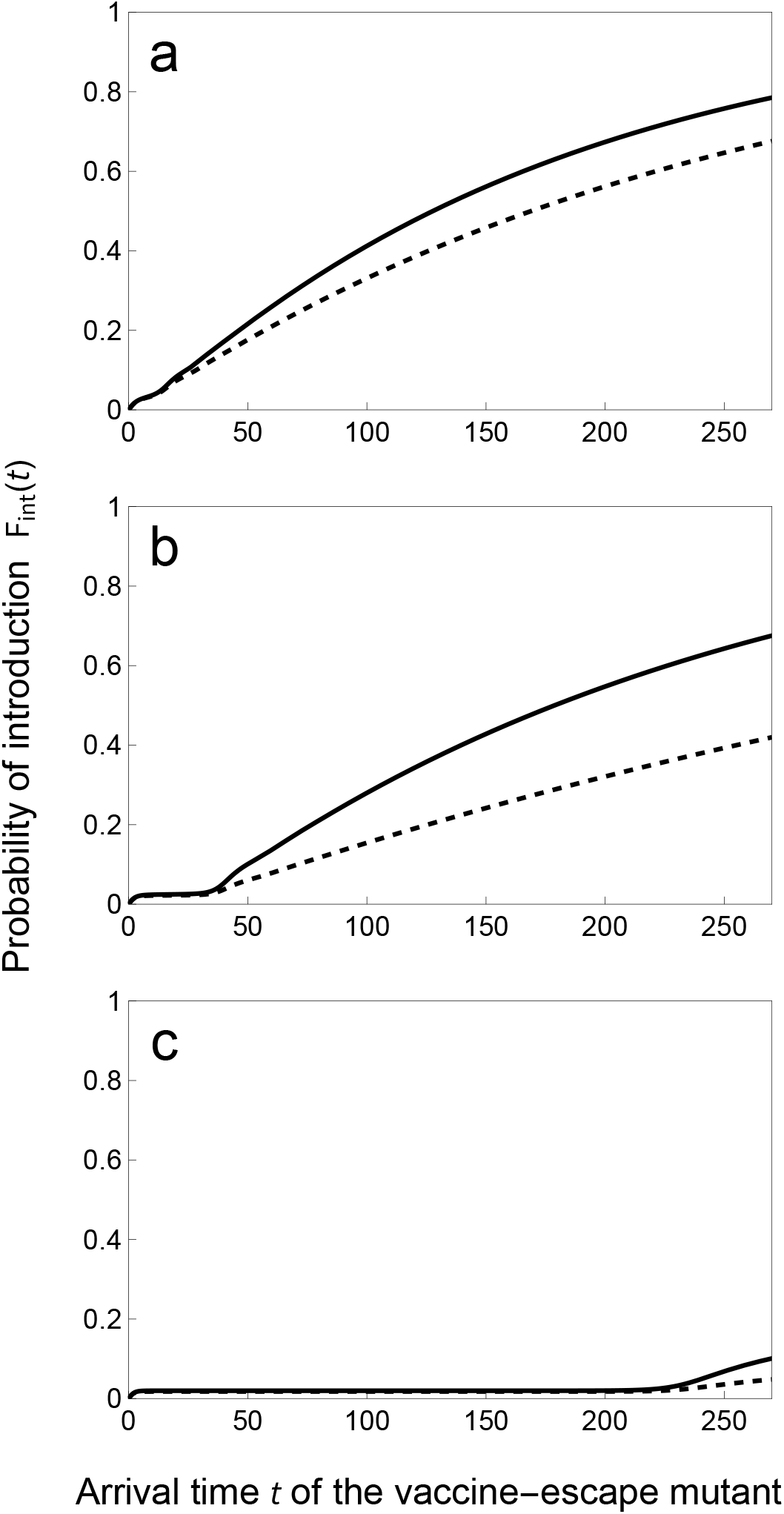
Faster vaccine rollout delays the arrival time of the first escape mutant. We plot the probability, *F*_int_(*t*), that the first escape mutant arrives prior to time *t* for different speeds of vaccination rollout: *υ* = 0.05 (top), 0.15 (middle) and 0.24 (bottom). We contrast a scenario where *θ*_v_ = *θ*_u_ (dashed line), and *θ*_v_ = 10 *× θ*_u_ (full line). Other parameter values: *θ*_u_ = 1, *ν* = *δ* = 3 10^−4^, *ω*_v_ = *ω*_r_ = 0.05, *α*_w_ = 0.02, *β*_w_ = 10, *γ*_w_ = 2, *ϵ*_w_ = 0.05, ℛ_w_ = 4.95. For these parameter values the critical rate of vaccination *υ*_c_ above which the wild-type pathogen is driven to extinction is *υ*_c_ ≈ 0.264 (see equation (8)).

#### 3.2.2 Step 2: Variant invasion

Immediately after its introduction, the dynamics of the vaccine-escape mutant may be approximated by a time-inhomogeneous birth-death process where the rate of birth (*i*.*e*., rate of new infections by the mutant) varies with the availability of susceptible hosts (see Section 3.1.2). The probability a mutant introduced at *T*_int_ = *t*_int_^1^ successfully invades (see [34] and Supplementary Information, §2) is:

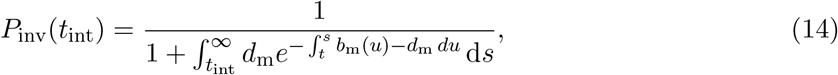

with *b*_m_(*t*) and *d*_m_ as defined above, (9), (10). In general, the integrals in (14) are impossible to compute exactly; in Methods, Section 5.2, we describe a fast numerical method.

Plotting the probability of invasion against the time of introduction, *t*, in **Figure 4** shows that the time at which the vaccine-escape mutant is introduced has a dramatic impact on the probability of escaping early extinction. If the mutant is introduced early, the density of susceptible vaccinated hosts remains very low and the selection for the vaccine-escape mutant is too small to prevent stochastic extinctions. The probability of invasion increases with selection, and thus with the density of vaccinated hosts, which tends to increase with time (see equation (6)).

**Figure 4:**
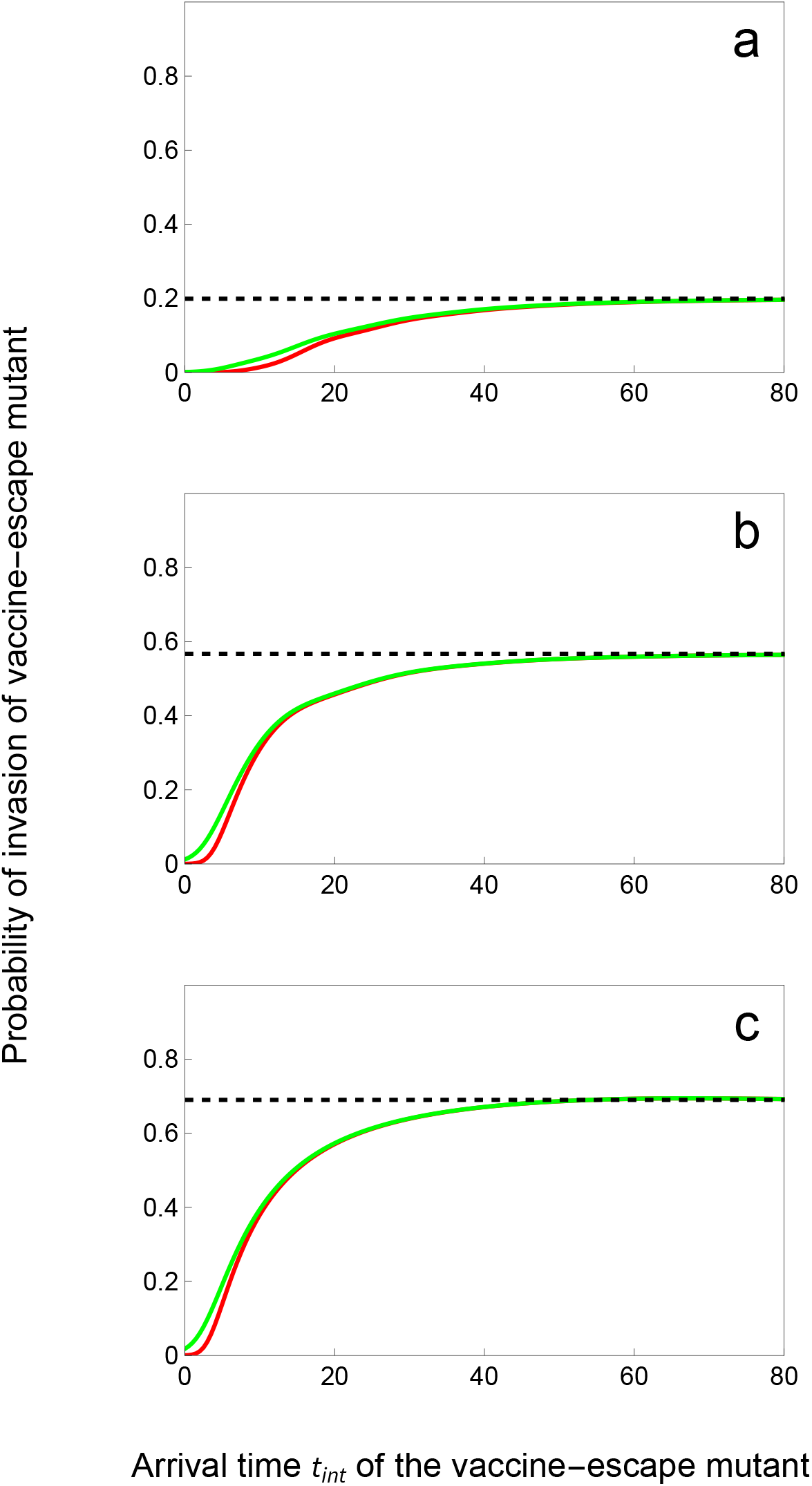
Probability of invasion of the vaccine-escape mutant increases with. *T*_int_. We plot the probability invasion *P*_inv_(*t*_int_) of a *slow* (green) and a *fast* (red) vaccine-escape mutant for different speeds of vaccination rollout: *υ* = 0.05 (top), 0.15 (middle) and 0.24 (bottom). The *slow* mutant: *α*_m_ = 0.02, *β*_m_ = 7, *γ*_m_ = 2, *ϵ*_m_ = 1, ℛ_m_ = 3.46. The *fast* mutant: *α*_m_ = 4.0606, *β*_m_ = 21, *γ*_m_ = 2, *ϵ*_m_ = 1, ℛ_m_ = 3.46. The probability of invasion 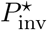 in the limit *t*_int_ → ∞ (see equation (16)) is indicated with the dashed black line. Other parameter values as in **Figure 3**: *ν* = *δ* = 3 10^−4^, *ω*_v_ = *ω*_r_ = 0.05, *α*_w_ = 0.02, *β*_w_ = 10, *γ*_w_ = 2, *ϵ*_w_ = 0.05, ℛ_w_ = 4.95.

Taking *t* → ∞ allows us to consider the situation when the vaccine-escape mutant appears at the post-vaccination endemic equilibrium, *i*.*e*., when the densities of unvaccinated and vaccinated susceptible hosts are 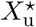 and 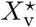, respectively (see Supplementary Information §1.3). At that point in time the effective per-generation reproduction ratio of genotype *i* (*i*.*e*., the expected number of secondary infections produced by pathogen genotype *i*) is (*cf*. (7)):

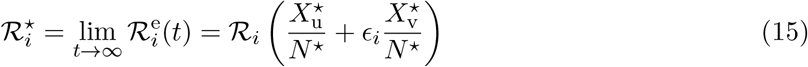

By definition, at the endemic equilibrium set by the wild-type pathogen we have 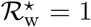. Hence, a necessary condition for the mutant to invade this equilibrium is 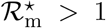, *i*.*e*., the effective reproduction number of the mutant has to be higher than that of the wild-type (see Supplementary Information, §1.3). However, this is not a sufficient condition: many mutants that satisfy this condition will rapidly go extinct due to demographic stochasticity. But in contrast to an early introduction of the mutant discussed above, the stochastic dynamics of the mutant is approximately a *time-homogeneous* branching process because the birth rate of the mutant approaches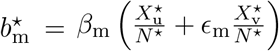. This birth rate is constant because the density of susceptible hosts remains constant at the endemic equilibrium. The probability of mutant invasion after introducing a single host infected by the mutant is thus (see Supplementary Information §3; **Figure 4**):

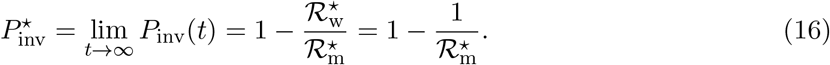

(note that we recover the strong-selection result of [49]). This expression shows that *at this endemic equilibrium* the fate of the mutant is fully governed by the per-generation reproduction ratio of the two strains, but does not depend on the specific values of the life-history traits of the mutant (provided the different vaccine-escape variants have the same value of 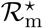).

Interestingly, unlike 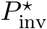, the probability *P*_inv_(*t*_int_) that a mutant introduced at time *T*_int_ = *t*_int_ successfully invades (14) is not governed solely by ℛ_*i*_, but rather depends on the life-history traits of the mutants. For instance, assume that two vaccine-escape mutants have the same values of ℛ_m_ and *ϵ*_m_ but they have very different life-history strategies. The “slow” strain has low rates of transmission and virulence (in green in **Figure 4**) while the “fast” strain has high rates of transmission and virulence (in red in **Figure 4**). **Figure 4** shows that the high mortality rate of hosts infected by the fast strain increases the risk of early extinction and lowers the probability of invasion relative to the slow strain. Hence, in the early stage of adaptation, pathogen life-history matters and favours slow strains with lower rates of transmission and virulence.

#### 3.2.3 Step 3: After variant invasion

Successful invasion of the vaccine-escape mutant means that it escaped the “danger zone” when its density is so low that it is very likely to go extinct (**Figure 1d-f**). After this invasion we can describe the dynamics of the polymorphic pathogen population using the deterministic approximation (4).

Because the invasion of the mutant at the endemic equilibrium set by the wild-type requires that 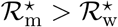, we might expect from the analysis of the deterministic model that the mutant would always replace the wild-type pathogen. That is, the wild-type pathogen would go extinct before the mutant (**Figure 1f**). This is indeed the case when the phenotypes of the mutant and the wild-type are not very different because of the “invasion implies fixation” principle [23, 9, 51]. Yet, this principle may be violated if the phenotype of the vaccine-escape mutant is very different than the phenotype of the wild-type.

First, the long-term coexistence of the two genotypes is possible (**Figure 1e**). The co-existence requires that each genotype is specialized on distinct types of host. The wildtype is specialised on unvaccinated hosts (*i*.*e*., ℛ_w_ *>* ℛ_m_) and the mutant is specialised on the vaccinated hosts (*i*.*e*., *ϵ*_m_ *> ϵ*_w_). Intermediate rates of vaccination maintain a mix of vaccinated and unvaccinated host wich promotes coexistence between the two genotypes (**Figure SI.2**). Second, the vaccine-escape mutant may be driven to extinction before the wild-type if its life-history traits induce massive epidemiological perturbations after its successful invasion (**Figure 1d**). As pointed out by previous studies, more transmissible and aggressive pathogen strategies may yield larger epidemics because the speed of the epidemic is governed by the per-capita growth rate *r*_*i*_, not by the per-generation reproduction ratio ℛ_*i*_ [19]. This explosive dynamics is driven by an over-exploitation of the host population and is immediately followed by a massive decline in the incidence of the vaccine-escape mutant. In a finite host population, this may result in the extinction of the vaccine-escape mutant before the wild-type [55]. We capture this outcome with a hybrid analytical-numerical approach that computes the probability *P*_fix_(*t*_int_) that the wild-type will go extinct before a mutant introduced at time *T*_int_ = *t*_int_ (see Methods, section 5.1). **Figure 5** shows that two vaccine-escape mutants may have very different probabilities of fixation, even if they have the same per-generation reproduction ratio. The numerical computation of the probability of fixation agrees very well with individual-based stochastic simulations. The faster strain is unlikely to go to fixation because invasion is followed by a period where the birth rate drops to very low levels (far below the mortality rates, **Figure SI.3**). In other words, a more aggressive strategy will more rapidly degrade its environment, by depleting susceptible hosts, which is known to increase the probability of extinction [10]. Interestingly, this effect is only apparent when the time of introduction, *T*_int_, is large. Indeed, when the mutant is introduced soon after the start of vaccination, its probability of invasion is already very low because its initial growth rate is negative (**Figure SI.3a, b, c**). When the mutant is introduced at intermediate times, the initial growth rate of the mutant is positive because some hosts are vaccinated (**Figure SI.3d, e, f**). If the vaccine-escape mutant is introduced later, the growth rate of the mutant is initially very high as many hosts are vaccinated (and thus susceptible to the vaccine-escape mutant) but this is rapidly followed by a drop in host density (especially pronounced with the faster strain) which prevents the long-term establishment of the faster strain (see **Figure SI.3g, h, i**).

**Figure 5:**
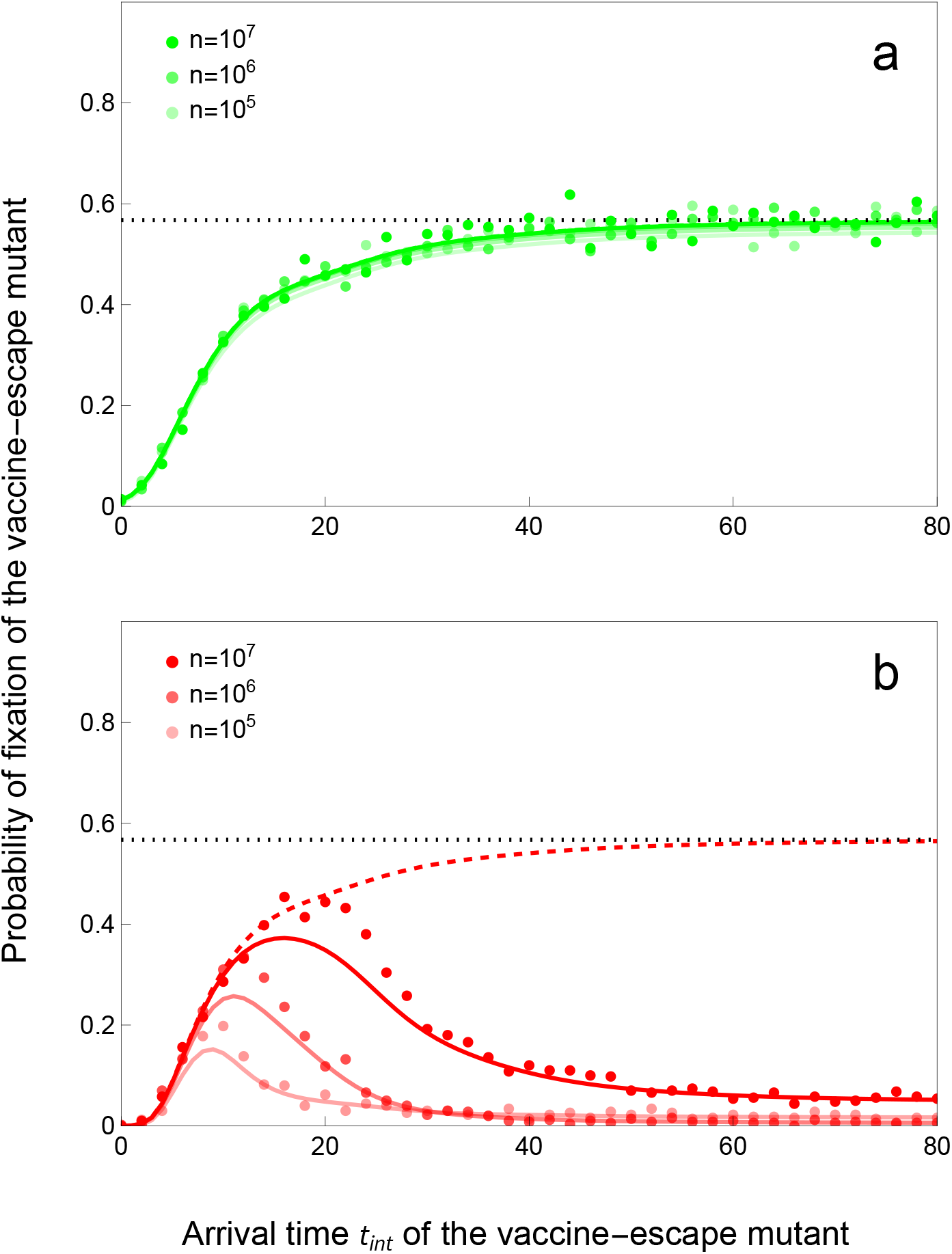
Probability of fixation of the vaccine-escape mutant may be low when *T*_int_ is large. We plot the probability of fixation of (A) a *slow* (green) and (B) a *fast* (red) vaccine-escape mutant for an intermediate speed of vaccination rollout: *υ* = 0.15. The *slow* mutant: *α*_m_ = 0.02, *β*_m_ = 7, *γ*_m_ = 2, *ϵ*_m_ = 1, ℛ_m_ = 3.46. The *fast* mutant: *α*_m_ = 4.0606, *β*_m_ = 21, *γ*_m_ = 2, *ϵ*_m_ = 1, ℛ_m_ = 3.46. The full colored lines give the probability of fixation *P*_fix_(*t*_inv_) computed numerically (see Methods section 5.4) and the dots give the results of individual-based simulations (see Methods section 5.6) for different values of *n* which affect the pathogen population size and the intensity of demographic stochasticity. We plot the probability of *invasion P*_inv_(*t*) (see **Figure 4**) with dashed colored line and its asymptotic value 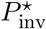 with a dotted black line. Other parameter values as in **Figure 3**: *ν* = *δ* = 3 10^−4^, *ω*_v_ = *ω*_r_ = 0.05, *p* = 0, *α*_w_ = 0.02, *β*_w_ = 10, *γ*_w_ = 2, *ϵ*_w_ = 0.05, ℛ_w_ = 4.95.

#### 3.2.4 The overall risk of pathogen adaptation

The overall probability that the pathogen will adapt to vaccination (*i*.*e*., that a vaccine-escape variant invades and eventually replaces or coexists with the wild-type) depends upon the probability that the mutation will arise (step 1) and the probability that this mutation will escape early extinction (step 2) and eventually go to fixation (step 3). It is particularly relevant to explore the effect of the speed of vaccination rollout on the overall probability that some vaccine-escape variant successfully invades at some time *T*_inv_ ≤ *t* after the start of the vaccination campaign (steps 1 and 2, **Figure 6**). Note that several variants can arise and fail to invade before finally a lucky variant manages to invade. We can use the probability of invasion *P*_inv_(*t*) of a variant introduced at time *t* to characterize the distribution, *F*_inv_(*t*), of the first time, *T*_inv_, at which a mutant is introduced that successfully invades. Using (12) and (14), this is

**Figure 6:**
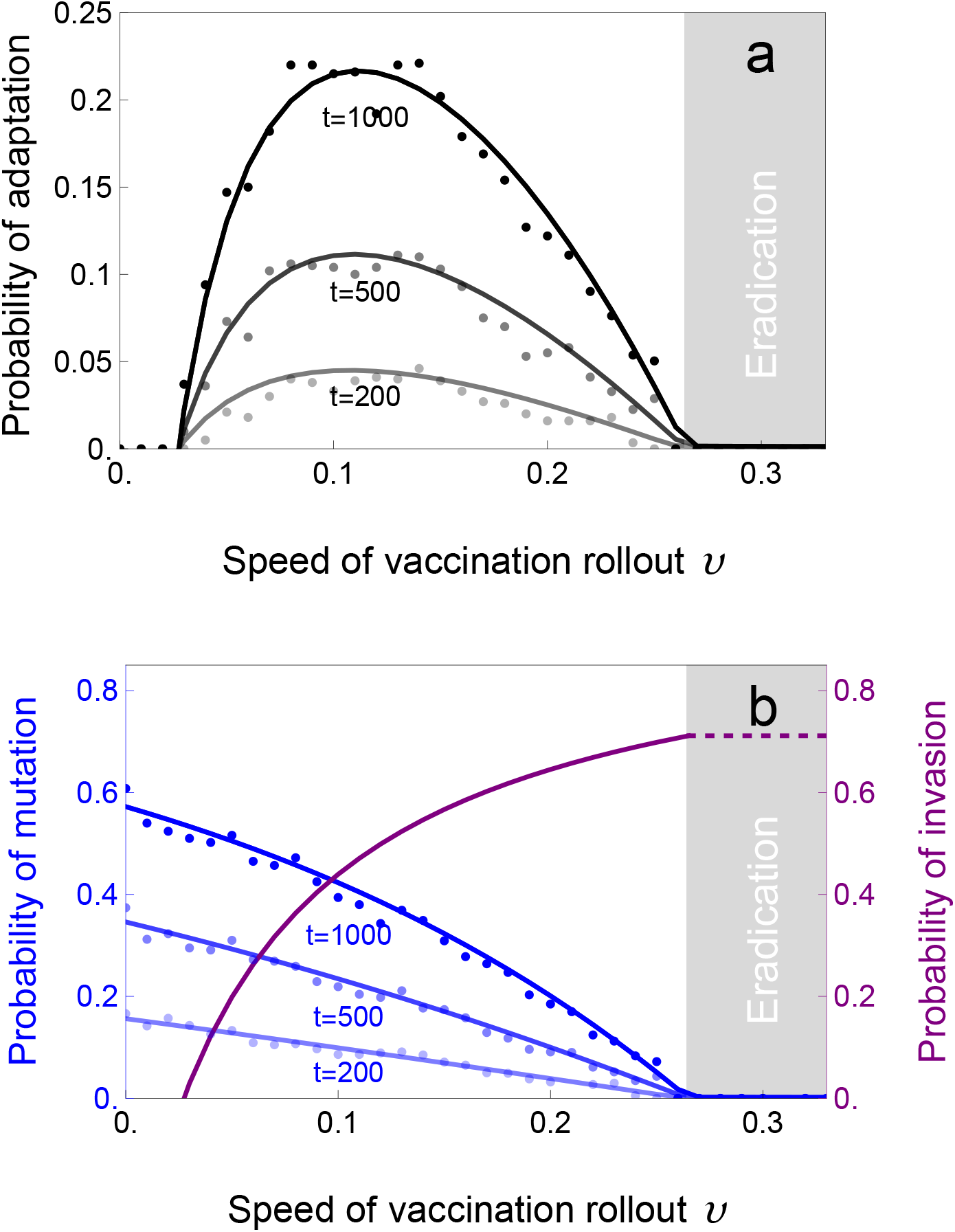
The probability of adaptation is maximised for intermediate speed of vaccination rollout. In (A) We plot the probability of adaptation *F*_inv_(*t*) (black lines) against the speed of vaccination rollout at different points in time. In (B) we plot the probability *F*_int_(*t*) of the introduction of at least one mutant before different points in time *t* (blue lines) and the probability 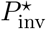 (purple line) which gives a good approximation of the probability of successful invasion of an escape-mutant. The dashed purple line gives the probability of invasion of the escape-mutant in the absence of the wild-type. The dots give the results of individual-based simulations (see Methods section 5.6). The vaccine-escape mutant is assumed to have the following phenotype (*slow* mutant in **Figure 4** and **5**): *α*_m_ = 0.02, *β*_m_ = 7, *γ*_m_ = 2, *ϵ*_m_ = 1, ℛ_m_ = 3.46. Other parameter values: *ν* = *δ* = 3 10^−4^, *n* = 10^6^, *ω*_v_ = *ω*_r_ = 0.05, *α*_w_ = 0.02, *β*_w_ = 10, *γ*_w_ = 2, *ϵ*_w_ = 0.05, ℛ_w_ = 4.95. The light gray area on the right-hand-side indicates the speed above which the wild-type pathogen is expected to be driven to extinction (*υ > υ*_c_ ≈ 0.264, see equation (8)).

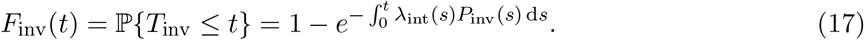

Compare (13) with (17) and note that the probability that no vaccine-escape mutant will ever *arise* is

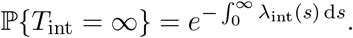

In contrast, the probability that no vaccine-escape mutant will ever *invade* is the larger probability

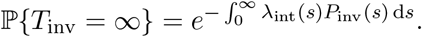

Note that *P*_inv_(*t*) converges as *t* → ∞ to 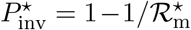 which is nonzero, so that ℙ*{T*_int_ = ∞*}* = 0 if and only if

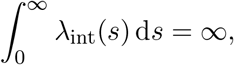

which in turn is true if and only if ℙ*{T*_inv_ = ∞*}* = 0. That is, the probability of adaptation is 1 if and only if *λ*_int_(*t*) is not integrable. In other words, the probability of adaptation is 1 in the limit *t* → ∞ when the wild-type is not driven to extinction by vaccination (*i*.*e*., *υ < υ*_c_) which implies that there is an uninterrupted flux of mutation producing vaccine-escape variants. One of these mutants will eventually escape extinction and invade. Yet, the time needed for a successful variant to appear may be very long (equation (17) and **Figure 6**).

When *υ > υ*_c_, vaccination is expected to eradicate the disease rapidly in our model (but see Supplementary Information §1.3). But an escape mutation may appear by mutation before eradication and rescue the pathogen population. This scenario fits squarely within the framework of classical “evolutionary rescue” modelling [43, 2, 7]. Yet, vaccination rollout is unlikely to be fast enough to eradicate the wildtype pathogen and, in this case, the probability of adaptation goes to 1 when *t* → ∞. Indeed, when *υ < υ*_c_, a vaccine-escape variant will eventually appear by mutation and invade. But what is less clear is how fast this adaptation will take place. We can use equation (17) to explore the effect of the speed of adaptation on the probability of pathogen adaptation at time *t* after the start of vaccination (*i*.*e*., the speed of adaptation). Crucially, the speed of pathogen adaptation is maximized for intermediate values of the speed of vaccination rollout. This is due to the antagonistic consequences the speed of the rollout has upon these two steps of adaptation (compare **Figures 3 and 4**). Faster rollout reduces *λ*_int_, the influx of new mutations, but increases *P*_inv_ because higher vaccination coverage yield stronger selection for vaccine-escape mutations. **Figure 6** illustrates how the speed of adaptation given in (17) results from the balance between the time-varying probability *F*_int_(*t*) that a variant is introduced by mutation before time *t* and the probability *P*_inv_(*t*) that this variant successfully invades (recall that 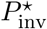 is a good approximation of this probability of invasion, see (16)).

## 4 Discussion

Vaccination is a powerful tool to control the spread of infectious diseases, but some pathogens evolve to escape the immunity triggered by vaccines (*e*.*g*., influenza, SARS-CoV-2). Will pathogens continue to adapt to the different vaccines that are being used to halt their spread? Does the likelihood of this adaptation depend on the speed of the vaccination rollout? To answer these questions we must first understand the different steps that may eventually lead to adaptation to vaccination.

Mutation is the fuel of evolution, and the first step of adaptation to vaccination is the mutational process that produces vaccine-escape variants. For instance, even if initial estimates of SARS-CoV-2 mutation rates were reassuringly low [52], the virus has managed to evolve higher rates of transmission [15, 62] and these adaptations are challenging control measures currently being used to slow down the ongoing pandemic. The ability of the new variants of SARS-CoV-2 to escape immunity is also worrying and indicates that viral adaption can weaken vaccine efficacy [63, 50]. The rate at which these potential vaccine-escape mutations are introduced depends on the density of hosts infected by the wild-type virus. In this respect, a faster rollout of vaccination is expected to delay the arrival of these mutations (**Figure 3**). Some authors, however, have argued that vaccine-escape mutations may arise more frequently in infected hosts which are partially immunized [56, 13, 16]. Our model can be used to explore the consequences of this within-host evolution in vaccinated hosts (*e*.*g*.,, taking *θ*_v_ *> θ*_u_). A larger value of *θ*_v_ increases the overall rate of mutation (**Figure 3**) but this effect is modulated by the fraction of the host population that is vaccinated. Consequently, when *θ*_v_ *> θ*_u_, the speed of vaccination rollout can have a non-monotonic effect on the probability that a vaccine-escape mutation is introduced (see **Figure SI.4**). Indeed, when the rate of vaccination remains low, the enhancing effect of vaccination on the rate of introduction of new mutations can counteract the delaying effect of faster vaccination rollout discussed above. But the probability that a vaccine-escape mutation is introduced drops to very low levels when the rate of vaccination gets closer to the critical vaccination rate *υ*_*c*_.

The second step of adaptation starts as soon as the vaccine-escape mutant has been introduced in the pathogen population. Will this new variant go extinct rapidly or will it start to invade? The answer to this question depends on the time at which the mutant is introduced. If the mutant is introduced when the population is not at an endemic equilibrium, the fate of the mutant depends on a time-varying birth rate which is driven by the fluctuations of the density of susceptible hosts. In our model, early introductions are likely to result in rapid extinction because there are simply not enough vaccinated hosts to favour the mutant over the wild-type. Moreover, we found that earlier introductions are likely to favour slower life-history strategies which are less prone to early extinction. If the introduction takes place later, when the system has reached a new endemic equilibrium, the fate of the mutant is solely governed by the effective per-generation ratio 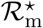 and does not depend on the life-history traits of the mutant. Slow and fast variants have equal probability to invade if they have the same 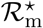. Altogether, our results suggest that earlier arrival may not always facilitate invasion since the probability of invasion is limited by the time-varying epidemiological state of the host population.

The third step of adaptation starts as soon as the hosts infected by the vaccine-escape mutant are abundant and the effect of demographic stochasticity on the dynamics of this mutation becomes negligible. Our analysis attempts to better characterize the dynamics of the mutant after invasion using a combination of deterministic and stochastic approximations. In principle, conditional on invasion, we can use the deterministic model (4) to describe the joint dynamics of the mutant and the wild-type. In particular, the speed at which the vaccine-escape mutant spreads in the pathogen population can be well approximated by the deterministic model. This may be particularly useful to address the impact of various vaccination strategies on the speed of the spread of a vaccine-escape variant [21]. In the present work we show that life-history traits of the vaccine-escape mutant drive the speed of its spread. Indeed, as pointed out before, the deterministic transient dynamics depends on the per-capita growth rate of the mutant *r*_m_, not its per-generation reproduction ratio ℛ_m_ [19]. Transient dynamics may favour a fast and aggressive variant (*i*.*e*., faster increase in frequency of this variant) because this life-history strategy may be more competitive away from the endemic equilibrium. Yet, this explosive strategy may be risky for the pathogen if it leads to epidemiological fluctuations that result in a massive drop in the number of infections. The consequences of such fluctuations on the extinction risk of the variant can be accounted for by a generalized birth-death process where the per-capita growth rate of the mutant varies with time. Epidemiological fluctuations lead to a degradation of the future environment (*i*.*e*., depletion of the density of susceptible hosts) which results in an increased risk of extinction [34, 10]. This effect has recently been analysed in a purely epidemiological model without vaccination [48]. In this simpler scenario, it is also relevant to make a distinction between early extinction (*i*.*e*., a *fizzle* in [48], **Figure 1c]**) and extinction after a successful invasion (*i*.*e*., an *epidemic burnout* in [48], **Figure 1d**) and it is possible to use a similar hybrid semi-deterministic approach to obtain accurate analytical approximations for both events.

A comprehensive understanding of pathogen dynamics after vaccination relies on the use of a combination of theoretical tools to capture the interplay between stochastic and deterministic forces. Here, we use a hybrid numerical-analytical approach to account for the three successive steps that may eventually lead to the fixation of a vaccine-escape mutant. This theoretical framework is particularly suitable to explore the influence of different vaccination strategies on the risk of pathogen adaptation. In particular, we show that this risk drops to very low levels even when the speed of vaccination rollout is below the threshold value that may eventually lead to eradication (*i*.*e*., *υ < υ*_c_). In other words, faster vaccination rollout makes sense even when eradication is infeasible, because faster rollout decreases both the number of cases and the likelihood of pathogen evolution. This conclusion is akin to the general prediction that the rate of pathogen adaptation should be maximized for intermediate immune pressure or for medium doses of chemotherapy at the within-host level [27, 53, 40, 1, 30, 17, 2]. Most of these earlier studies focused on evolutionary rescue scenarios where the wild-type is expected to be rapidly driven to extinction by human intervention. Our versatile theoretical framework, however, allows us to deal with a broader range of situations where the intervention is not expected to eradicate the wild-type pathogen. Accounting for the dynamics of the wild-type affects both the flux of mutation and the fate of these mutations. Note how our decomposition of the factors acting on the probability of adaptation (**Figure 6**) provides a validation of the verbal argument often used in earlier studies to explain the higher rate of pathogen adaptation for intermediate levels of vaccination coverage of drug concentration [27, 53, 40, 56].

The framework we have developed can be readily extended to explore many other situations. For instance, our model can be modified to explore the influence of temporal variations in the environment that could be driven by seasonality or by non-pharmaceutical interventions (NPIs). We explored a situation where the transmission rate of all variants is periodically reduced by a quantity 1 − *c*(*t*), where *c*(*t*) is a measure of the intensity of NPIs. These periodic interventions affect both the flux of mutations and the probability that these mutations invade. In particular, NPIs lower the probability of mutant introduction through the reduction in the density of hosts infected by the wild-type (**Figure SI.4**). As a consequence, the probability of adaptation is reduced when vaccination is combined with periodic control measures. Hence, our approach helps to understand the interaction between vaccination and NPI discussed in earlier studies [54, 42].

We have made several simplifying assumptions that need to be relaxed to confidently apply our findings to a broader range of pathogens such as the current SARS-CoV-2 pandemic (see section 5.6 in the Methods). First, one should study situations where the pathogen population has not reached an endemic equilibrium when vaccination starts to be applied. We carried out additional simulations showing that starting the vaccination rollout sooner (*i*.*e*., just after the start of the epidemic) tends to promote the probability of invasion of the escape mutant (**Figure SI.5**). Indeed, at the onset of the epidemic the density of susceptible hosts is higher (*i*.*e*., the birth rate of the infection is high relative to the endemic equilibrium) and the risk of early extinction of the mutant is reduced. Second, it is important to relax the assumption that natural immunity is perfect. We carried out additional simulations showing that when naturally immune hosts, like vaccinated hosts, can be reinfected the probability of invasion of the escape mutant increases (**Figure SI.6**). This effect is particularly strong just after the start of vaccination. Indeed, if naturally immune hosts are equivalent to vaccinated hosts, selection to escape immunity is present even before the start of vaccination and one may thus expect the speed of adaptation to be much faster. Yet, the vaccination strategy can affect the rate of adaptation. In particular, we find that faster rates of vaccination always reduce the rate of adaptation via the reduction of the influx of escape mutants (**Figure SI.7**). Another important extension of our model would be to study the effect of a diversity of vaccines in the host population. We did not explore this effect in the present study but this diversity of immune profiles among vaccinated hosts could slow down pathogen adaptation if the escape of different vaccines requires distinct mutations [60, 11, 46].

Finally, it is important to recall that we focus here on a simplified scenario where we analyse the evolutionary epidemiology of an isolated population. In real-life situations the arrival time may depend more on the immigration of new variants from abroad than on local vaccination policies. The influence of migration remains to be investigated in spatially structured models where vaccination may vary among populations [24].

## 5 Methods

In this section, we present how extinction, invasion and fixation probabilities may be obtained under strong-selection assumptions when a mutant strain appears in a host-pathogen system that is away from its endemic equilibrium. Our essential tools are the deterministic ordinary differential equations (Section 3.1.1) and birth-and-death process approximations, (Section 3.1.2). The former allows us to consider the situation when all strains are abundant, the latter when at least one strain is rare. We will limit ourselves to an informal treatment, presenting heuristic arguments and deferring rigorous proofs and sharp error bounds to a future treatment.

### 5.1 Approximating the Fixation Probability

Suppose that the mutant strain introduced at time *T*_int_ = *t*_int_ successfully invades; we next consider the probability *P*_fix_(*t*_int_) that the mutant will outcompete the wild-type and go to fixation. Fixation of the mutant occurs if it is still present when the wild-type strain disappears. If we let 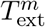 and 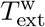 be the extinction times of mutant and wild-type strains, the probability of mutant fixation is thus 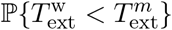 which we may decompose as

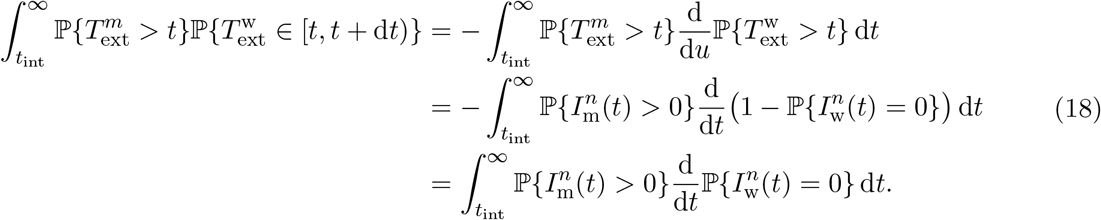

We obtain estimates of 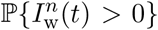 and 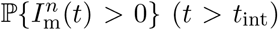 by now approximating *both* mutant *and* wild-type strains by birth-death-processes *Ĩ*_m_(*t*) and *Ĩ*_w_(*t*) (see Section 3.1.2).

The birth rates for the two types, *i* = w, m, are given by

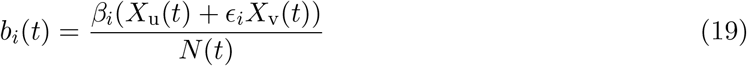

and the death rates are

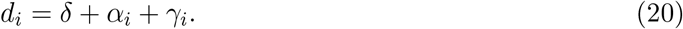

for *i* ∈ *{*w, m*}*.

As previously, we are approximating the frequency of unvaccinated and vaccinated hosts by their deterministic approximations

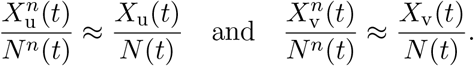

and compute the latter using the ordinary differential equations (4). Unlike previously, when we assumed that the mutant was rare, and took *Y*_m_(*t*) ≡ 0, we are now allowing the possibility that the mutant is abundant, and cannot neglect the effect of the mutant strain on *X*_v_ and *X*_u_. In particular, we need to take care in choosing the initial conditions of (4) to account for the fact that we consider the time of appearance of the first mutant that successfully invades and so are conditioning on the non-extinction of the mutant strain, and to account for the inherent variability in the time required to invade; this results in a random initial condition for the deterministic dynamics (see Supplementary Information §4 for details). In practice, we find that the randomness has negligible effect, but we must still take the conditioning into account. To do so, we first use (4) with *Y*_m_(0) = 0 (so *Y*_m_(*t*) ≡ 0 for *t >* 0) and initial conditions (SI.1) to compute the epidemiological dynamics of the wild-type from time 0 up until the the introduction of the mutant at time *t*_int_. Then, at time *t*_int_, we restart (4) with new initial conditions: we use the values *X*_u_(*t*_int_), *X*_v_(*t*_int_), *N* (*t*_int_) and *Y*_w_(*t*_int_) computed assuming *Y*_m_(0) = 0, and take

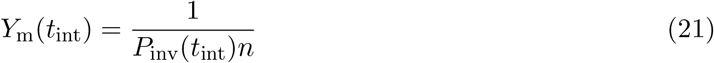

(see Supplementary Information §4 for details). Crucially, the initial density of the mutant depends on the probability of successful invasion of the mutant *P*_inv_(*t*_int_) obtained above (14).

Provided we use (4) with the appropriate initial conditions as previously, the birth rates of both the wild-type and mutant strains are approximately deterministic, and from [34], we have:

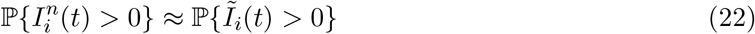

Under the branching assumption, the lines of descent of distinct infected individuals are independent, hence the probability that strain *i* vanishes by time *t* is the product of the probabilities that each line of descent vanishes,

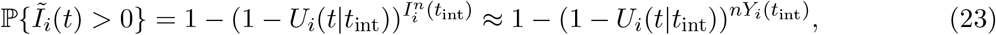

where

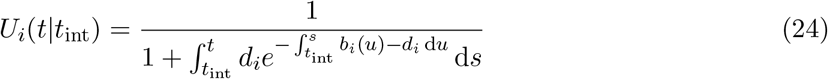

is the probability that an individual infected with strain *i* ∈ *{*w, m*}* present at time *t*_int_ has descendants alive at time *t > t*_int_ and we approximate the initial *number* of individuals infected with strain *i* using the *frequencies* obtained using (4) and (21):

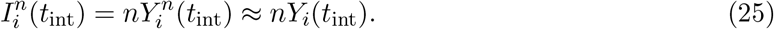

Below in Section 5.2.3, we give a fast numerical method for computing *U*_*i*_(*t*|*t*_int_).

### 5.2 Auxiliary Functions

In the following we present a simple, yet versatile, hybrid (*i*.*e*., semi-deterministic and semi-numerical) framework which allows us to approximate the probabilities associated with different steps of adaptation (mutation, invasion, fixation) by adding auxiliary equations describing stochastic phenomena to the deterministic ordinary differential equations describing the global population dynamics.

#### 5.2.1 Introduction of the variant by mutation (step 1)

Recall 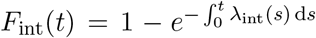, (13), where *λ*_int_ (*t*) = *θ*_u_ *Y*_uw_ (*t*) + *θ*_v_ *Y*_vw_ (*t*), (12). Rather than computing the integral – which would require that we compute *λ*_int_(*s*) (and thus *Y*_uw_(*s*) and *Y*_vw_(*s*)) for every *s < t*, we observe that the cumulative hazard 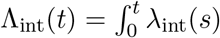 d*s* can be computed by combining (4) with initial conditions (SI.1) and the *auxilliary* differential equation

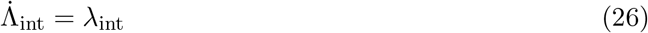

with initial condition Λ_int_(0) = 0. The use of this auxiliary equation reduces computational effort by obtaining Λ_int_(*t*) simultaneously with *Y*_uw_(*t*) and *Y*_vw_(*t*) (as opposed to computing the latter two and then integrating).

#### 5.2.2 Invasion of the variant (step 2)

In practice, the probability of mutant invasion (14) involves integrals that cannot be explicitly computed, and we must compute it numerically. To do so, we make use of of one of the steps involved in computing *P*_inv_(*t*_int_) in [34]. There, it is shown that

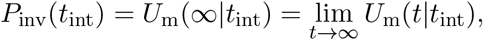

where

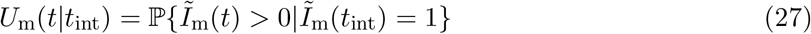

is obtained via a pair of auxiliary functions

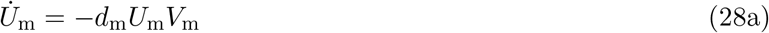

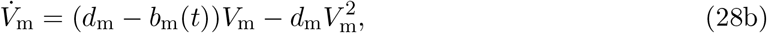

with initial conditions

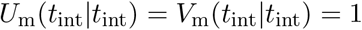

(*N*.*B*., 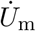 and 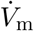 denote the derivatives with respect to *t*). We compute *b*_m_(*t*), which depends on *X*_u_(*t*), *X*_v_(*t*) and *N* (*t*) (see (10)), via (4). In practice, we cannot compute *U*_m_(∞|*t*); to obtain an approximation we approximate it by *U*_m_(*t*|*t*_int_) for the first *t* sufficiently large that

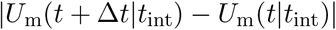

is less than our desired threshold of error, where Δ*t* is the step size in our numerical scheme.

#### 5.2.3 Fixation of the variant (step 3)

In practice, we need two pairs of auxiliary equations, *i* ∈ *{*w, m*}*, to track the probabilities that some descendant of a wild-type or mutant individual that was present at *t*_int_ is still alive at time *t*:

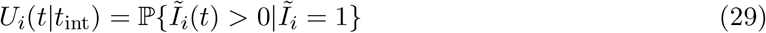

Exactly as in (28) above, these satisfy

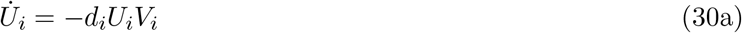

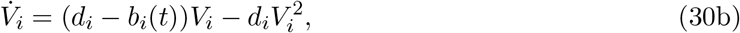

with

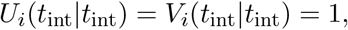

for *i* ∈ *{*u, v*}*.

To compute the probability of fixation, we first consider the probability that fixation occurs prior to time *t*, which is derived in exactly the same manner as (18).

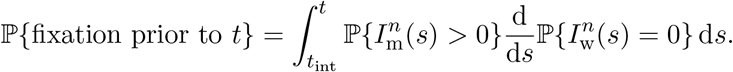

Proceeding as in Section 5.1, approximating the probabilities 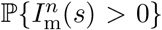 and 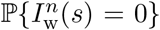 by *{Ĩ*_m_(*s*) *>* 0*}* and P*{Ĩ*_w_(*s*) = 0*}* and initial number of hosts infected with the wild-type using the deterministic density, 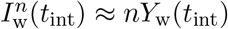, using the branching property (23) this is approximately

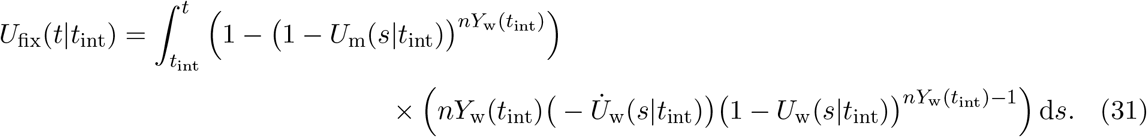

Differentiating yields the following auxiliary equation for *U*_fix_(*t*):

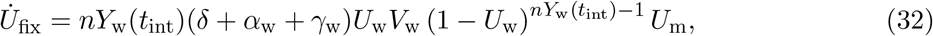

with initial condition *U*_fix_(*t*_int_|*t*_int_) = 0. We estimate the fixation probability as

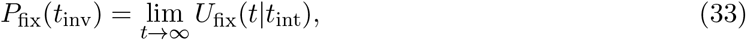

approximating the limit at infinity as we did for *P*_inv_(*t*_int_) in Section 5.2.2above.

#### 5.2.4 The overall risk of pathogen adaptation

We numerically compute the cumulative density function *F*_inv_(*t*) = ℙ*{T*_inv_ ≤ *t}* of the first arrival time *T*_inv_ of a vaccine-escape mutant that successfully invades (17) analogously to *F*_int_ (Section 5.2.1), using the auxiliary equation

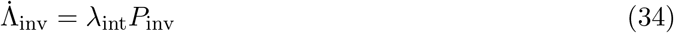

with initial condition Λ_inv_(0) = 0, computing *Y*_uw_(*t*) and *Y*_vw_(*t*) – and thus *λ*_int_(*t*) – using (4) with initial conditions (SI.1).

### 5.3 Stochastic simulations

We carried out stochastic simulations to check the validity of our results. We developed an individual-based simulation program for the Markov process described in **Table 1** and using the parameter values given in **Table SI.1**. In order to match the assumption used in our analysis we start the simulation when the system is at its endemic equilibrium before vaccination. Then we introduce a single host infected with the mutant pathogen at a time *t*_int_ after the start of vaccination and we let the simulation run until one of the pathogen variants (the wild-type or the mutant) goes extinct. If the wild-type goes extinct first we record this run as a “mutant fixation event”. We ran 1000 replicates for each set of parameters and we plot the proportion of runs that led to mutant fixation in **Figure 5**. We also used our simulations to confirm our prediction on the speed of viral adaptation in **Figure 6**. In this scenario we allowed the vaccine-escape variant to be introduced by mutation from the wild-type genotype. We carried out 1000 simulations and monitored (i) the frequency of the escape mutant at different points in time after the start of vaccination (**Figure 6a**) (ii) the number of introduction events by mutation and (**Figure 6b**). We also used this simulation approach to go beyond the scenarios used in our analysis to check the robustness of some of our results.

## Data Availability

This is a theoretical study.

## Data accessibility

The simulation code used to carry out stochastic simulations has been deposited on zenodo 10.5281/zenodo.12655541.

## Acknowledgements

SG acknowledges support from the CNRS PEPS 2022 grant “VaxDurable”.

## Footnotes

1 To clarify, *T*_int_ is the random time at which a mutation arises; when we specify *T*_int_ = *t*_int_, we are conditioning on the event in which the random quantity *T*_int_ takes the fixed value *t*_int_.

