## Supplementary Information for "The speed of vaccination rollout and the risk of pathogen adaptation"

### 1 Equilibria and Invasion Fitnesses

Below, we obtain and provide notation for the various equilibria of (4) that are relevant to our study. All equilibria correspond to a 5-tuple of densities $\big(X_{\mathrm{u}}, X_{\mathrm{v}}, Y_{\mathrm{uw}}, Y_{\mathrm{um}}, Z\big)$.

#### 1.1 Pre-Vaccination Endemic Equilibrium

We first assume that prior to the start of vaccination at time $t = 0$ the wild-type pathogen reached its endemic equilibrium whereas no mutant stain is present (but see §5 below), which (since it will be the initial condition for (4) for all computations in the Main Text) we denote $\big(X_{\mathrm{u}}(0), 0, Y_{\mathrm{uw}}(0), 0, Z(0)\big)$. Setting all derivatives equal to zero and $X_{\mathrm{v}} = Y_{\mathrm{vw}} = 0$ in (4) we find a unique solution,

$$X_{\mathrm{u}}(0) = \frac{\nu(\delta + \gamma_{\mathrm{w}} + \omega_{\mathrm{r}})}{\delta(\beta_{\mathrm{w}} - \alpha_{\mathrm{w}}) + \omega_{\mathrm{r}}(\mathcal{R}_{\mathrm{w}}(\alpha_{\mathrm{w}} + \delta) - \alpha_{\mathrm{w}})}, \tag{SI.1a}$$

$$Y_{\mathrm{uw}}(0) = \frac{\nu(\mathcal{R}_{\mathrm{w}} - 1)(\delta + \omega_{\mathrm{r}})}{\delta(\beta_{\mathrm{w}} - \alpha_{\mathrm{w}}) + \omega_{\mathrm{r}}(\mathcal{R}_{\mathrm{w}}(\alpha_{\mathrm{w}} + \delta) - \alpha_{\mathrm{w}})}, \tag{SI.1b}$$

and

$$Z(0) = \frac{\nu(\mathcal{R}_{\mathrm{w}} - 1)\gamma_{\mathrm{w}}}{\delta(\beta_{\mathrm{w}} - \alpha_{\mathrm{w}}) + \omega_{\mathrm{r}}(\mathcal{R}_{\mathrm{w}}(\alpha_{\mathrm{w}} + \delta) - \alpha_{\mathrm{w}})}. \tag{SI.1c}$$

#### 1.2 Disease Free Equilibrium

We next consider the possible equilibria in the presence of vaccination but in the absence of the pathogen. We will denote such a disease free equilibrium by $(X_{\mathrm{u}}^{\varnothing}, X_{\mathrm{v}}^{\varnothing}, 0, 0, 0)$, with $N^{\varnothing} = X_{\mathrm{u}}^{\varnothing} + X_{\mathrm{v}}^{\varnothing}$.

Setting $Y_{\mathrm{uw}} = Y_{\mathrm{um}} = Y_{\mathrm{vw}} = Y_{\mathrm{vm}} = Z = 0$ and solving $\dot{X}_{\mathrm{u}} = \dot{X}_{\mathrm{v}} = 0$ in (4) for the remaining variables yields the unique-long term equilibrium in the absence of infection,

$$X_{\mathrm{u}}^{\varnothing} = \frac{\nu(\delta + \omega_{\mathrm{v}})}{\delta(\delta + \upsilon + \omega_{\mathrm{v}})}, \tag{SI.2a}$$

$$X_{\mathrm{v}}^{\varnothing} = \frac{\upsilon\nu}{\delta(\delta + \upsilon + \omega_{\mathrm{v}})}, \tag{SI.2b}$$

and $N^{\varnothing} = X_{\rm u}^{\varnothing} + X_{\rm v}^{\varnothing} = \nu/\delta$.

The wild-type strain can invade this disease free equilibrium if and only its effective reproduction number (7) at this equilibrium exceeds one: $\mathcal{R}_{\rm w}^{\varnothing} > 1$, where

$$\mathcal{R}_{\rm w}^{\varnothing} = \mathcal{R}_{\rm w}\left(\frac{X_{\rm u}^{\varnothing}}{N^{\varnothing}} + \epsilon_{\rm w}\frac{X_{\rm v}^{\varnothing}}{N^{\varnothing}}\right) = \mathcal{R}_{\rm w}\frac{\delta + \epsilon_{\rm w}\upsilon + \omega_{\rm v}}{\delta + \upsilon + \omega_{\rm v}} < \mathcal{R}_{\rm w}. \tag{SI.3}$$

Equivalently, $\mathcal{R}_{\rm w}^{\varnothing} < 1$ gives us a criterion for the vaccination intensity required to prevent the emergence of a pathogen in a community of epidemiologically naïve individuals: $\upsilon > \upsilon_{\rm c}$ with

$$\upsilon_{\rm c} = \frac{(\mathcal{R}_{\rm w} - 1)(\delta + \omega_{\rm v})}{1 - \mathcal{R}_{\rm w}\epsilon_{\rm w}}. \tag{SI.4}$$

Note how the critical rate of vaccination $\upsilon_{\rm c}$ becomes arbitrarily large as $\epsilon_{\rm w}\mathcal{R}_{\rm w} \to 1$.

##### 1.3 Endemic Equilibria and Backwards Bifurcations

We next characterize the endemic equilibria, when the wild-type strain is fixed in the pathogen population. We denote such an equilbrium by $(X_{\rm u}^{\star}, X_{\rm v}^{\star}, Y_{\rm uw}^{\star}, Y_{\rm vw}^{\star}, Z^{*})$ with $N^{\star} = X_{\rm u}^{\star} + X_{\rm v}^{\star} + S_{\rm r}^{\star} + Y_{\rm uw}^{\star} + Y_{\rm vw}^{\star}$.

Setting $Y_{\rm um} = Y_{\rm vm} = 0$ in $\dot{X}_{\rm u} = \dot{X}_{\rm v} = 0$ in (4) and solving for $X_{\rm u}$ and $X_{\rm v}$ yields

$$\begin{aligned} X_{\rm u}^{\star} &= \frac{(\nu + \omega_{\rm r}Z^{\star})}{\mathcal{D}}\left(\epsilon_{\rm w}\beta_{\rm w}\frac{Y_{\rm w}^{\star}}{N^{\star}} + \delta + \omega_{\rm v}\right) \\ X_{\rm v}^{\star} &= \frac{(\nu + \omega_{\rm r}Z^{\star})}{\mathcal{D}}\upsilon, \end{aligned} \tag{SI.5}$$

with

$$\mathcal{D} = \left(\delta + \upsilon + \beta_{\rm w}\frac{Y_{\rm w}^{\star}}{N^{\star}}\right)\left(\delta + \omega_{\rm v} + \epsilon_{\rm w}\beta_{\rm w}\frac{Y_{\rm w}^{\star}}{N^{\star}}\right) - \omega_{\rm v}\upsilon, \tag{SI.6}$$

Setting $Y_{\rm um} = Y_{\rm vm} = 0$ in $\dot{Z} = 0$ in (4) and solving for $Z$ yields

$$Z^{\star} = \frac{\gamma_{\rm w}Y_{\rm w}^{\star}}{\delta + \omega_{\rm r}}, \tag{SI.7}$$

We then have

$$N^\star = X_\mathrm{u}^\star + X_\mathrm{v}^\star + Y_\mathrm{w}^\star + Z^\star = \frac{\nu - \alpha_\mathrm{w} Y_\mathrm{w}^\star}{\delta}. \tag{SI.8}$$

Setting $Y_\mathrm{um} = Y_\mathrm{vm} = 0$ in $\dot{Y}_\mathrm{w} = 0$, we find that at any endemic equilibrium, we must have

$$\mathcal{R}_\mathrm{w}\left(\frac{X_\mathrm{u}^\star}{N^\star} + \epsilon_\mathrm{w}\frac{X_\mathrm{v}^\star}{N^\star}\right) = 1, \tag{SI.9}$$

Substituting (SI.5), (SI.7), and (SI.8) into (SI.9) gives us a quadratic polynomial that must be satisfied by any non-zero solution $Y_\mathrm{w}^\star$. It turns out, however, that one obtains a polynomial that is much more readily interpreted [2] by instead solving for the equilibrium force of infection,

$$h_\mathrm{w}^\star = \frac{\beta_\mathrm{w} Y_\mathrm{w}^\star}{N^\star}. \tag{SI.10}$$

One then has

$$A(h_\mathrm{w}^\star)^2 + Bh_\mathrm{w}^\star + C = 0, \tag{SI.11}$$

where

$$A = \epsilon_\mathrm{w}(\gamma_\mathrm{w} + \delta + \omega_\mathrm{r}), \tag{SI.12a}$$

$$B = \epsilon_\mathrm{w}(\delta + \gamma_\mathrm{w} + \omega_\mathrm{r})\upsilon + (\delta + \omega_\mathrm{r})(\delta + \gamma_\mathrm{w} + \omega_\mathrm{r}) - \epsilon_\mathrm{w}(\delta + \omega_\mathrm{r})(\delta + \alpha_\mathrm{w} + \gamma_\mathrm{w})(\mathcal{R}_\mathrm{w} - 1), \tag{SI.12b}$$

and

$$C = (\delta + \omega_\mathrm{r})(\delta + \upsilon + \omega_\mathrm{v})(\delta + \gamma_\mathrm{w} + \alpha_\mathrm{w})(1 - \mathcal{R}_\mathrm{w}^\varnothing). \tag{SI.12c}$$

Recall that (SI.11) has real roots if $\Delta = B^2 - 4AC > 0$, which are given by the quadratic formula, $Y_\mathrm{w}^\star = \frac{-B \pm \sqrt{\Delta}}{2A}$. In particular, if $C < 0$ – which occurs if and only if $\mathcal{R}_\mathrm{w}^\varnothing > 1$ – then, since $A > 0$, $\Delta > B^2$ and there is a unique positive root, $\frac{-B+\sqrt{\Delta}}{2A}$. As observed above, if $\mathcal{R}_\mathrm{w}^\varnothing > 1$ then the wild-type can invade the disease free equilibrium.

When $C > 0$ ($\mathcal{R}_{\mathrm{w}}^{\varnothing} < 1$), there are three possibilities:

(i) if $\Delta < 0$, then (SI.11) has no real roots, and there is no endemic equilibrium;

(ii) if $\Delta > 0$ and $B > 0$, then, since $\Delta < B^2$, both roots $\frac{-B \pm \sqrt{\Delta}}{2A}$ have non-zero complex parts, and again there is no endemic equilbrium;

(iii) if $\Delta > 0$ and $B < 0$, then (SI.11) has two positive roots.

The latter scenario leads to a phenomenon known as a *backwards bifurcation* (see [6] for a simple example in a model with vaccination; [3] surveys the phenomenon more generally in epidemic models): while the wild-type cannot invade the disease free equilibrium, if initially present in sufficient abundance, it can persist indefinitely despite vaccination reducing the effective reproduction number of the wild-type, $\mathcal{R}_{\mathrm{w}}^{\mathrm{e}}$, below one. This is because the smaller root, $\frac{-B-\sqrt{\Delta}}{2A}$, is an unstable equilibrium that divides the basin of attraction of zero from the basin of attraction of $\frac{-B+\sqrt{\Delta}}{2A}$.

Given $h_{\mathrm{w}}^{\star}$, one can use (SI.10) and (SI.8) to obtain $Y_{\mathrm{w}}^{\star} = \frac{\nu h_{\mathrm{w}}^{\star}}{\beta_{\mathrm{w}}\delta + \alpha_{\mathrm{w}} h_{\mathrm{w}}^{\star}}$, which in turn give us $X_{\mathrm{u}}^{\star}$, $X_{\mathrm{v}}^{\star}$, $Z^{\star}$ and $N^{\star}$ via equations (SI.5) and (SI.6), (SI.7), and (SI.8) respectively. Finally, setting $\dot{Y}_{\mathrm{uw}} = 0$ and $\dot{Y}_{\mathrm{vw}} = 0$ in (4) allow us to solve for $Y_{\mathrm{uw}}^{\star}$ and $Y_{\mathrm{vw}}^{\star}$:

$$
\begin{aligned}
Y_{\mathrm{uw}}^{\star} &= \frac{\beta_{\mathrm{w}} Y_{\mathrm{w}}^{\star} X_{\mathrm{u}}^{\star}}{N^{\star}(\delta + \alpha_{\mathrm{w}} + \gamma_{\mathrm{w}})} = \mathcal{R}_{\mathrm{w}} \frac{X_{\mathrm{u}}^{\star}}{N^{\star}} Y_{\mathrm{w}}^{\star} \\
Y_{\mathrm{vw}}^{\star} &= \frac{\epsilon_{\mathrm{w}} \beta_{\mathrm{w}} Y_{\mathrm{w}}^{\star} X_{\mathrm{v}}^{\star}}{N^{\star}(\delta + \alpha_{\mathrm{w}} + \gamma_{\mathrm{w}})} = \epsilon_{\mathrm{w}} \mathcal{R}_{\mathrm{w}} \frac{X_{\mathrm{v}}^{\star}}{N^{\star}} Y_{\mathrm{w}}^{\star}.
\end{aligned}
$$

##### 1.3.1 Backwards Bifurcations and Pathogen Extinction

If we wish to use $\mathcal{R}_{\mathrm{w}}^{\varnothing} < 1$ as a criterion for pathogen eradication, we need to exclude a backwards bifurcation. Since, by assumption, $\mathcal{R}_{\mathrm{w}} > 1$, we cannot conclude *a priori* from (SI.12b) that $B > 0$. However, observe that the coefficient of $\upsilon$ in (SI.12b) is always positive, so $B$ will always be positive if

$$
\epsilon(\delta + \alpha_{\mathrm{w}} + \gamma_{\mathrm{w}})(\delta + \omega_{\mathrm{r}})(\mathcal{R}_{\mathrm{w}} - 1) \leq (\delta + \omega_{\mathrm{v}})(\delta + \gamma_{\mathrm{w}} + \omega_{\mathrm{r}}),
$$

*i.e.*, provided

$$\alpha_{\mathrm{w}} \leq \alpha_{\mathrm{c}} = \frac{(\delta+\omega_{\mathrm{v}})(\delta+\gamma_{\mathrm{w}}+\omega_{\mathrm{r}}) - \epsilon(\mathcal{R}_{\mathrm{w}}-1)(\delta+\gamma_{\mathrm{w}})(\delta+\omega_{\mathrm{r}})}{\epsilon(\delta+\omega_{\mathrm{r}})(\mathcal{R}_{\mathrm{w}}-1)}. \tag{SI.13}$$

Since $\omega_{\mathrm{r}} \leq \omega_{\mathrm{v}}$, and, by assumption $\epsilon_{\mathrm{w}}\mathcal{R}_{\mathrm{w}} < 1$,

$$\begin{aligned}(\delta+\omega_{\mathrm{v}})(\delta+\gamma_{\mathrm{w}}+\omega_{\mathrm{r}}) &- \epsilon(\mathcal{R}_{\mathrm{w}}-1)(\delta+\gamma_{\mathrm{w}})(\delta+\omega_{\mathrm{r}}) \\ &\geq (\delta+\omega_{\mathrm{r}})\big((\delta+\gamma_{\mathrm{w}}+\omega_{\mathrm{r}}) - \epsilon(\mathcal{R}_{\mathrm{w}}-1)(\delta+\gamma_{\mathrm{w}})\big) \\ &> (\delta+\omega_{\mathrm{r}})(\delta+\gamma_{\mathrm{w}})(1-\epsilon\mathcal{R}_{\mathrm{w}}+\epsilon) > 0.\end{aligned}$$

Thus, $\alpha_{\mathrm{c}} > 0$ and (SI.13) is satisfied for sufficiently small $\alpha_{\mathrm{w}} \geq 0$. For instance, using the default parameter values used in all our numerical examples (see Table **??**) we have $\alpha_{\mathrm{w}} = 0.02 < \alpha_{\mathrm{c}} = 8.38$. In other words, backward bifurcation is not feasible in our numerical examples and pathogen eradication is expected to occur when the rate of vaccination is above the critical vaccination rate given by (8).

More generally, we can write (SI.12b) as

$$B = \epsilon_{\mathrm{w}}(\delta+\gamma_{\mathrm{w}}+\omega_{\mathrm{r}})\upsilon + \epsilon_{\mathrm{w}}(\delta+\omega_{\mathrm{r}})(\mathcal{R}_{\mathrm{w}}-1)(\alpha_{\mathrm{c}}-\alpha_{\mathrm{w}}),$$

so that even if $\alpha_{\mathrm{w}} > \alpha_{\mathrm{c}}$, a backwards bifurcation can still be excluded ($B > 0$) with a sufficiently high rate of vaccination, $\upsilon > \upsilon_{\mathrm{bb}}$, where

$$\upsilon_{\mathrm{bb}} = \frac{(\delta+\omega_{\mathrm{r}})(\mathcal{R}_{\mathrm{w}}-1)(\alpha_{\mathrm{w}}-\alpha_{\mathrm{c}})}{(\delta+\gamma_{\mathrm{w}}+\omega_{\mathrm{r}})}. \tag{SI.14}$$

#### 2 Time-Inhomogeneous Birth and Death Processes

We briefly summarize the results we use from [5]:

**Theorem 1** ([5]). *Let $Z(t)$ be a continuous-time linear birth-death process with time-varying birth* *rate $b(t)$ and death rate $d(t)$, i.e., such that*

$$\mathbb{P}\left\{Z(t+h)=k+1 \mid Z(t)=k\right\}=b(t)kh+o(h)$$

$$\mathbb{P}\left\{Z(t+h)=k-1 \mid Z(t)=k\right\}=d(t)kh+o(h)$$

*Then, $Z(t)$ has mean*

$$m_k(t|s)=\mathbb{E}[Z(t)|Z(s)=k]=ke^{\int_0^t b(u)-d(u)\,\mathrm{d}u} \tag{SI.15}$$

*and probability generating function*

$$F_k(x,t|s)=\mathbb{E}\big[x^{Z(t)}\big|Z(s)=k\big]=F_1(x,t|s)^k, \tag{SI.16}$$

*where*

$$F_1(x,t|s):=\mathbb{E}\big[x^{Z(t)}\big|Z(s)=1\big]=\frac{q(t|s)+\big(1-q(t|s)-\eta(t|s)\big)x}{1-\eta(t|s)x}, \tag{SI.17}$$

*for*

$$\eta(t|s)=\frac{\int_s^t e^{\int_s^r b(u)-d(u)\,\mathrm{d}u}b(r)\,\mathrm{d}r}{1+\int_s^t e^{\int_s^r b(u)-d(u)\,\mathrm{d}u}b(r)\,\mathrm{d}r}, \tag{SI.18}$$

*and*

$$q(t|s):=\mathbb{P}\{Z(t)=0|Z(s)=1\}=\frac{\int_s^t e^{-\int_s^r b(u)-d(u)\,\mathrm{d}u}d(r)\,\mathrm{d}r}{1+\int_s^t e^{-\int_s^r b(u)-d(u)\,\mathrm{d}u}d(r)\,\mathrm{d}r}. \tag{SI.19}$$

*In particular,*

$$\mathbb{P}\left\{Z(t)>0|Z(s)=1\right\}=\frac{1}{1+\int_s^t e^{-\int_s^r b(u)-d(u)\,\mathrm{d}u}d(r)\,\mathrm{d}r},$$

*and the probability of extinction in finite time, given $Z(s)=1$ is*

$$q(s)=\lim_{t\to\infty}q(t|s)=\frac{J(s)}{1+J(s)}, \tag{SI.20}$$

*which is equal to 1 if and only if the integral*

$$J(s):=\int_s^\infty e^{-\int_s^r b(u)-d(u)\,\mathrm{d}u}d(r)\,\mathrm{d}r \tag{SI.21}$$

*diverges. More generally, if $Z(s) = k$ then the probability of extinction is $q(s)^k$.*

In Section 3.2.2, we apply Theorem 1 to the problem of pathogen invasion by taking $s$ to be the time of introduction of a novel mutant, $t_{\text{int}}$, $b(t) = b_{\text{m}}(t)$ (10), and $d(t) = d_{\text{m}}$ (9). In Section 5.1, we apply it to pathogen fixation by again taking $s$ to be $t_{\text{int}}$, $b(t) = b_i(t)$ (19), and $d(t) = d_i$ (20) for $i \in \{\text{w}, \text{m}\}$.

#### 3 Invasion at the post-vaccination endemic equilibrium

Here, we show that, as one might expect, the extinction and invasion probabilities are continuous in the limit as $t_{\text{int}} \to \infty$ (*cf.* (16)).

**Proposition 1.** *Let $Q_{\text{inv}}(t_{\text{int}}) = 1 - P_{\text{inv}}(t_{\text{int}})$, where $P_{\text{inv}}(t_{\text{int}})$ is given by (14). Then,*

$$\lim_{t_{\text{int}} \to \infty} Q_{\text{inv}}(t_{\text{int}}) = \frac{1}{\mathcal{R}_{\text{m}}^{\star}}.$$

*Proof.* First, observe that

$$\begin{aligned} J(t_{\text{int}}) &= \int_{t_{\text{int}}}^{\infty} e^{-\int_{t_{\text{int}}}^{t} \frac{\beta_{\text{m}}(X_{\text{u}}(u) + \epsilon_{\text{m}} X_{\text{v}}(u))}{N(u)} - (\delta + \alpha_{\text{m}} + \gamma_{\text{m}})\,\text{d}u} (\delta + \alpha_{\text{m}} + \gamma_{\text{m}})\,\text{d}t \\ &= \int_{0}^{\infty} e^{-\int_{0}^{t} \frac{\beta_{\text{m}}(X_{\text{u}}(u+t_{\text{int}}) + \epsilon_{\text{m}} X_{\text{v}}(u+t_{\text{int}}))}{N(u+t_{\text{int}})} - (\delta + \alpha_{\text{m}} + \gamma_{\text{m}})\,\text{d}u} (\delta + \alpha_{\text{m}} + \gamma_{\text{m}})\,\text{d}t. \end{aligned}$$

Choose $\varepsilon > 0$ sufficiently small that $\frac{\beta_{\text{m}} X_{\text{u}}^{\star} + \epsilon_{\text{m}} \beta_{\text{m}} X_{\text{v}}^{\star}}{N^{\star}} - \varepsilon > \delta + \alpha_{\text{m}} + \gamma_{\text{m}}$. Since $(X_{\text{u}}(t), X_{\text{v}}(t), N(t)) \to (X_{\text{u}}^{\star}, X_{\text{v}}^{\star}, N^{\star})$ as $t \to \infty$, there exists $t_{\varepsilon} > 0$ such that

$$\left| \frac{\beta_{\text{m}}(X_{\text{u}}(t) + \epsilon_{\text{m}} X_{\text{v}}(t))}{N(t)} - \frac{\beta_{\text{m}}(X_{\text{u}}^{\star} + \epsilon_{\text{m}} X_{\text{v}}^{\star})}{N^{\star}} \right| < \varepsilon.$$

Thus, for $t_{\text{int}} > t_{\varepsilon}$, we have

$$\begin{aligned} &\int_{0}^{t} \frac{\beta_{\text{m}}(X_{\text{u}}(u+t_{\text{int}}) + \epsilon_{\text{m}} X_{\text{v}}(u+t_{\text{int}}))}{N(u+t_{\text{int}})} - (\delta + \alpha_{\text{m}} + \gamma_{\text{m}})\,\text{d}u \\ &\qquad > \left( \frac{\beta_{\text{m}}(X_{\text{u}}^{\star} + \epsilon_{\text{m}} X_{\text{v}}^{\star})}{N^{\star}} - \varepsilon - (\delta + \alpha_{\text{m}} + \gamma_{\text{m}}) \right) t. \end{aligned}$$

Now, by Birkhoff's ergodic theorem, for every $t_{\mathrm{int}}$,

$$\lim_{t\to\infty}\frac{1}{t}\int_0^t \frac{\beta_{\mathrm{m}}(X_{\mathrm{u}}(u+t_{\mathrm{int}})+\epsilon_{\mathrm{m}}X_{\mathrm{v}}(u+t_{\mathrm{int}}))}{N(u+t_{\mathrm{int}})}\,\mathrm{d}u = \frac{\beta_{\mathrm{m}}(X_{\mathrm{u}}^{\star}+\epsilon_{\mathrm{m}}X_{\mathrm{v}}^{\star})}{N^{\star}}.$$

Thus, there exists $t_{\varepsilon,t_{\mathrm{int}}}$ such that

$$\left|\frac{1}{t}\int_0^t \frac{\beta_{\mathrm{m}}(X_{\mathrm{u}}(u+t_{\mathrm{int}})+\epsilon_{\mathrm{m}}X_{\mathrm{v}}(u+t_{\mathrm{int}}))}{N(u+t_{\mathrm{int}})}\,\mathrm{d}u - \frac{\beta_{\mathrm{m}}(X_{\mathrm{u}}^{\star}+\epsilon_{\mathrm{m}}X_{\mathrm{v}}^{\star})}{N^{\star}}\right| < \frac{\varepsilon}{2}.$$

Further, since $\frac{1}{t}\int_0^t \frac{\beta_{\mathrm{m}}(X_{\mathrm{u}}(u+t_{\mathrm{int}})+\epsilon_{\mathrm{m}}X_{\mathrm{v}}(u+t_{\mathrm{int}}))}{N(u+t_{\mathrm{int}})}\,\mathrm{d}u$ is continuous in $t_{\mathrm{int}}$, there exists $\delta_{t_{\mathrm{int}}}>0$ such that

$$\left|\frac{1}{t}\int_0^t \frac{\beta_{\mathrm{m}}(X_{\mathrm{u}}(u+t_{\mathrm{int}})+\epsilon_{\mathrm{m}}X_{\mathrm{v}}(u+t_{\mathrm{int}}))}{N(u+t_{\mathrm{int}})}\,\mathrm{d}u - \frac{1}{t}\int_0^t \frac{\beta_{\mathrm{m}}(X_{\mathrm{u}}(u+s)+\epsilon_{\mathrm{m}}X_{\mathrm{v}}(u+s))}{N(u+s)}\,\mathrm{d}u\right| < \frac{\varepsilon}{2}$$

for all $|s-t_{\mathrm{int}}|<\delta_{t_{\mathrm{int}}}$. By compactness of the closed interval, $\{B_{\delta_{t_{\mathrm{int}}}}(t_{\mathrm{int}}) : t_{\mathrm{int}}\in[0,t_\varepsilon]\}$ admits a finite subcover $\{B_{\delta_{t_1}}(t_1),\ldots,B_{\delta_{t_K}}(t_K)\}$. Set

$$t'_\varepsilon = \max_k t_{\varepsilon,t_k}.$$

Then, for all $t_{\mathrm{int}}\in[0,t_\varepsilon]$ and all $t>t'_\varepsilon$, we have $t_{\mathrm{int}}\in B_{\delta_{t_k}}(t_k)$ for some $k$ and $t>t_{\varepsilon,t_k}$, whence

$$\frac{1}{t}\int_0^t \frac{\beta_{\mathrm{m}}(X_{\mathrm{u}}(u+t_{\mathrm{int}})+\epsilon_{\mathrm{m}}X_{\mathrm{v}}(u+t_{\mathrm{int}}))}{N(u+t_{\mathrm{int}})}\,\mathrm{d}u > \frac{\beta_{\mathrm{m}}(X_{\mathrm{u}}^{\star}+\epsilon_{\mathrm{m}}X_{\mathrm{v}}^{\star})}{N^{\star}} - \varepsilon.$$

By compactness of the domain and continuity, we then have that

$$C_\varepsilon = \sup_{t_{\mathrm{int}}\le t_\varepsilon}\sup_{t\le t'_\varepsilon} e^{-\int_0^t \frac{\beta_{\mathrm{m}}(X_{\mathrm{u}}(u+t_{\mathrm{int}})+\epsilon_{\mathrm{m}}X_{\mathrm{v}}(u+t_{\mathrm{int}}))}{N(u)}\,\mathrm{d}u + \left(\frac{\beta_{\mathrm{m}}(X_{\mathrm{u}}^{\star}+\epsilon_{\mathrm{m}}X_{\mathrm{v}}^{\star})}{N^{\star}}+\varepsilon\right)t}$$

is finite, whereas

$$e^{-\int_0^t \frac{\beta_{\mathrm{m}}(X_{\mathrm{u}}(u+t_{\mathrm{int}})+\epsilon_{\mathrm{m}}X_{\mathrm{v}}(u+t_{\mathrm{int}}))}{N(u+t_{\mathrm{int}})}-(\delta+\alpha_{\mathrm{m}}+\gamma_{\mathrm{m}})\,\mathrm{d}u} \le C_\varepsilon e^{-\left(\frac{\beta_{\mathrm{m}}(X_{\mathrm{u}}^{\star}+\epsilon_{\mathrm{m}}Xv^{\star})}{N^{\star}}-\varepsilon-(\delta+\alpha_{\mathrm{m}}+\gamma_{\mathrm{m}})\right)t}$$

for all $t_{\mathrm{int}}$ and $t$. The latter function is integrable on $[0,\infty)$, and we may thus apply Lebesgue's dominated convergence theorem (see *e.g.,* [10, Theorem 4.16, p. 91]) to interchange limit and integral, so

$$\begin{aligned}
\lim_{t_{\mathrm{int}}\to\infty} J(t_{\mathrm{int}}) &= \lim_{t_{\mathrm{int}}\to\infty} \int_0^\infty e^{-\int_0^t \frac{\beta_{\mathrm{m}}(X_{\mathrm{u}}(u+t_{\mathrm{int}})+\epsilon_{\mathrm{m}} X_{\mathrm{v}}(u+t_{\mathrm{int}}))}{N(u+t_{\mathrm{int}})} - (\delta+\alpha_{\mathrm{m}}+\gamma_{\mathrm{m}})\,\mathrm{d}u} (\delta+\alpha_{\mathrm{m}}+\gamma_{\mathrm{m}})\,\mathrm{d}t \\
&= \int_0^\infty e^{-\left(\frac{\beta_{\mathrm{m}}(X_{\mathrm{u}}^\star+\epsilon_{\mathrm{m}} X_{\mathrm{v}}^\star)}{N^\star} - (\delta+\alpha_{\mathrm{m}}+\gamma_{\mathrm{m}})\right)t} (\delta+\alpha_{\mathrm{m}}+\gamma_{\mathrm{m}})\,\mathrm{d}t \\
&= \frac{1}{\frac{\mathcal{R}_{\mathrm{m}}(X_{\mathrm{u}}^\star+\epsilon_{\mathrm{m}} X_{\mathrm{v}}^\star)}{N^\star} - 1}.
\end{aligned}$$

The result follows. □

### 4 Bridging the Invasion and Fixation Phases

Previously, we computed the invasion probability by ignoring the effect of the rare mutant strain on $(X_{\mathrm{u}}(t), X_{\mathrm{v}}(t), N(t))$, with (4), setting $Y_{\mathrm{m}}(t)$ identically equal to 0 for all $t$ (see Main Text, §3.2.2). Once the mutant is abundant, however, we can no longer neglect its effect on the abundance of susceptible hosts. Unfortunately, the full stochastic model is intractable. Instead, we approximate the effect of the mutant strain on the host density by the deterministic approximation (4) with $Y_{\mathrm{m}}(t) > 0$. To do so, we must first make an appropriate choice for the initial value $Y_{\mathrm{m}}$.

We will say that the mutant strain has *successfully invaded* when it has achieved a positive density of $\varepsilon$, that is, when there are $\varepsilon n$ individuals infected with the mutant strain, for a fixed $\varepsilon > 0$ that is small and independent of $n$. From time $t_{\mathrm{int}}$ until the first time when the mutant population size exceeds $\varepsilon n$, say $T_\varepsilon$, we can again neglect the effect of the rare mutant strain on host abundance.

After time $T_\varepsilon$, the macroscopic presence of mutants has a non-negligible effect on the dynamics of $(X_{\mathrm{u}}(t), X_{\mathrm{v}}(t), N(t))$ (which in turn determine the probability of late mutant extinction). We could account for this by re-starting (4) at $T_\varepsilon$ with $Y_{\mathrm{m}}(T_\varepsilon) = \varepsilon$. One difficulty with this approach is that $T_\varepsilon$ is random, depending on the initial size fluctuations of the mutant population. Furthermore, there is no an *a priori* choice for $\varepsilon$ (see *e.g.,* [7, §2.2] for a detailed rigorous treatment of how invasion probabilities depend on initial mutant frequency in the SIS model).

Instead of estimating $T_\varepsilon$, we use the fact that $T_\varepsilon - t_{\text{int}}$ is of the order of a constant times $\ln(n)$ for large $n$ (see *e.g.*, [8, Supplementary Information §8.2.2]), and approximate $Y_{\text{m}}(t)$ by its large-time behaviour to get an "effective initial condition" at time $t_{\text{int}}$ that generates the same trajectory as if we took the initial condition $Y_{\text{m}}(T_\varepsilon) = \varepsilon$.

To do this, we shall need the following corollary to Theorem 1:

**Corollary 1.** *Let $Z(t)$ be a continuous time linear birth-death process with time-varying birth rate $b(t)$ and death rate $d(t)$ such that $Z(s) = 0$, and suppose that*

$$\lim_{t\to\infty} e^{\int_s^t b(u)-d(u)\,\mathrm{d}u} = \infty.$$

*Set*

$$W_s(t) = e^{-\int_s^t b(u)-d(u)\,\mathrm{d}u} Z(t)$$

*then, as $t \to \infty$, $W_s(t)$ converges a.s. to a random variable $W_s$ on $[0, \infty)$. Moreover, $\mathbb{P}\{W_s = 0\} = q(s)$, whereas*

$$\mathbb{P}\{W_s > x | W_s > 0\} = e^{-(1-q(s))x},$$

*where $q(s)$ is the probability that $Z(t) = 0$ for some $t > 0$, (SI.20).*

*Proof.* We begin by observing that $W(t)$ is positive, $\mathbb{E}[W(t)] = 1$ for all $t$, and

$$\begin{aligned}
\mathbb{E}[W(t)|Z(r)] &= \mathbb{E}\left[e^{-\int_s^t b(u)-d(u)\,\mathrm{d}u} Z(t) \middle| Z(r)\right] \\
&= e^{-\int_s^t b(u)-d(u)\,\mathrm{d}u} e^{\int_r^t b(u)-d(u)\,\mathrm{d}u} Z(r) \\
&= e^{-\int_s^r b(u)-d(u)\,\mathrm{d}u} Z(r) \\
&= W_s(r),
\end{aligned}$$

so $W_s(t)$ is a martingale. Almost sure convergence to a limit $W_s$ then follows from Doob's martingale convergence theorem (see *e.g.*, [9, Theorem 10, p. 8]).

To characterize the distribution of $W$, recall the probability generating function for $Z(t)$,

$F_1(x,t|s)$, given by (SI.16) in Theorem 1. The Laplace transform for $W(t) = e^{-\int_0^t b(u)-d(u)\,\mathrm{d}u} Z(t)$ is

$$\begin{aligned}
L_W(\theta,t|s) &= \mathbb{E}[e^{-\theta W(t)}|Z(s)=1] \\
&= F_1(e^{-e^{-\int_s^t b(u)-d(u)\,\mathrm{d}u}\theta}, t|s) \\
&= \frac{q(t|s) + \big(1-q(t|s)-\eta(t|s)\big)e^{-e^{-\int_s^t b(u)-d(u)\,\mathrm{d}u}\theta}}{1-\eta(|s)e^{-e^{-\int_s^t b(u)-d(u)\,\mathrm{d}u}\theta}} \\
&= \frac{\big(1-q(t|s)-\eta(t|)\big)e^{\int_s^t b(u)-d(u)\,\mathrm{d}u}\left(e^{-e^{-\int_s^t b(u)-d(u)\,\mathrm{d}u}\theta}-1\right) + e^{\int_s^t b(u)-d(u)\,\mathrm{d}u}(1-\eta(t|s))}{e^{\int_s^t b(u)-d(u)\,\mathrm{d}u}\big(1-\eta(t|s)\big) - \eta(t|s)e^{\int_s^t b(u)-d(u)\,\mathrm{d}u}\left(e^{-e^{-\int_s^t b(u)-d(u)\,\mathrm{d}u}\theta}-1\right)}
\end{aligned}$$

Now, by assumption,

$$\lim_{t\to\infty} e^{-\int_s^t b(u)-d(u)\,\mathrm{d}u} = 0,$$

whereas from (SI.18),

$$\lim_{t\to\infty} \eta(t|s) = 1,$$

so, integrating by parts, and recalling (SI.19),

$$\begin{aligned}
e^{\int_s^t b(u)-d(u)\,\mathrm{d}u}\big(1-\eta(t|s)\big) &= \frac{1}{e^{-\int_s^t b(u)-d(u)\,\mathrm{d}u} + \int_s^t e^{-\int_s^r b(u)-d(u)\,\mathrm{d}u} b(r)\,\mathrm{d}r} \\
&= \frac{1}{1+\int_s^t e^{-\int_s^r b(u)-d(u)\,\mathrm{d}u} d(r)\,\mathrm{d}r} \\
&= 1-q(t|s).
\end{aligned}$$

Thus,

$$\lim_{t\to\infty} L_W(\theta,t|s) = \frac{q(s)\theta + 1 - q(s)}{1-q(s)+\theta} = q(s) + (1-q(s))\frac{1-q(s)}{1-q(s)+\theta}.$$

This is the Laplace transform of

$$q(s)\delta_0(w) + (1-q(s))^2 e^{-(1-q(s))w},$$

where $\delta_0(w)$ is a Dirac mass at zero. *i.e.* the distribution of a random variable that is 0 with probability $q(s)$ and, conditionally upon being non-zero, is exponentially distributed with mean $\frac{1}{1-q(s)}$. By Lévy's continuity theorem (see *e.g.,* [1, Theorem 3.3.6]), the limit $W_s$ has this distribution. □

To apply this to the invasion of a mutant strain, let $\tilde{I}_{\mathrm{m}}(t)$ $(t > t_{\mathrm{int}})$ be a birth-and-death process with rates $\frac{\beta_{\mathrm{m}}(X_{\mathrm{u}}(t)+\epsilon_{\mathrm{m}} X_{\mathrm{v}}(t))}{N(t)}$ and $\delta + \alpha_{\mathrm{m}} + \gamma_{\mathrm{m}}$, respectively, and $\tilde{I}_{\mathrm{m}}(t_{\mathrm{int}}) = 1$, and let $Y_{\mathrm{m}}(\varepsilon, \cdot)$ be the solution to (4) with initial condition $Y_{\mathrm{m}}(\varepsilon, T_\varepsilon) = \varepsilon$. Applying the corollary to $\tilde{I}_{\mathrm{m}}(t)$ yields a random variable $W_{t_{\mathrm{int}}}$ such that $\mathbb{P}\{W_{t_{\mathrm{int}}} = 0\} = Q_{\mathrm{inv}}(t_{\mathrm{int}})$:

$$e^{-\int_{t_{\mathrm{int}}}^{T_\varepsilon} \frac{\beta_{\mathrm{m}}(X_{\mathrm{u}}(u)+\epsilon_{\mathrm{m}} X_{\mathrm{v}}(u))}{N(u)} - (\delta+\alpha_{\mathrm{m}}+\gamma_{\mathrm{m}})\,\mathrm{d}u} \varepsilon n = e^{-\int_{t_{\mathrm{int}}}^{T_\varepsilon} \frac{\beta_{\mathrm{m}}(X_{\mathrm{u}}(u)+\epsilon_{\mathrm{m}} X_{\mathrm{v}}(u))}{N(u)} - (\delta+\alpha_{\mathrm{m}}+\gamma_{\mathrm{m}})\,\mathrm{d}u} \tilde{I}_{\mathrm{m}}(\varepsilon, T_\varepsilon) \to W_{t_{\mathrm{int}}}$$

as $n \to \infty$, since $T_\varepsilon = \inf\{t > t_{\mathrm{int}} : \tilde{I}_{\mathrm{m}}(t) \geq \varepsilon n\} \to \infty$ as $n \to \infty$.

While this can, in principle, be solved for $T_\varepsilon$, we instead observe that

$$\begin{aligned}
\varepsilon &= Y_{\mathrm{m}}(\varepsilon, T_\varepsilon) \\
&= Y_{\mathrm{m}}(\varepsilon, t) e^{\int_t^{T_\varepsilon} \frac{\beta_{\mathrm{m}}(X_{\mathrm{u}}(u)+\epsilon_{\mathrm{m}} X_{\mathrm{v}}(u))}{N(u)} - (\delta+\alpha_{\mathrm{m}}+\gamma_{\mathrm{m}})\,\mathrm{d}u} \\
&\approx Y_{\mathrm{m}}(\varepsilon, t) \frac{\varepsilon n}{W_t},
\end{aligned}$$

whence, for $n \gg 1$, we have $Y_{\mathrm{m}}(\varepsilon, t) \approx \frac{W_t}{n}$.

Thus with probability $Q_{\mathrm{inv}}(t_{\mathrm{int}})$, $W_{t_{\mathrm{int}}} = 0$ and thus $Y_{\mathrm{m}}(\varepsilon, t_{\mathrm{int}}) = 0$ and $Y_{\mathrm{m}}(\varepsilon, t) \equiv 0$ for all $t \geq t_{\mathrm{int}}$, corresponding to extinction of the birth-and-death process. On the other hand, conditioned on non-extinction, the initial condition is exponentially distributed with mean

$$\mathbb{E}\left[\frac{W_{t_{\mathrm{int}}}}{n} \middle| W_{t_{\mathrm{int}}} > 0\right] = \frac{1}{(1 - Q(t_{\mathrm{int}}))n} = \frac{1}{P_{inv}(t_{\mathrm{int}})n} > \frac{1}{n}.$$

In practice, this means that the trajectories $Y_{\mathrm{m}}(t)$ with initial conditions $Y_{\mathrm{m}}(T_\varepsilon) = \varepsilon$ and $Y_{\mathrm{m}}(t_{\mathrm{int}}) = \frac{W_{t_{\mathrm{int}}}}{n}$ coincide, and we may integrate (4) starting directly from time $t_{\mathrm{int}}$, sampling the initial condition $Y_{\mathrm{m}}(t_{\mathrm{int}}) = \frac{W_t}{n}$, from an exponential distribution. Each such trajectory corresponds to a different sample path of the random process $I^n_{\mathrm{m}}(t)$, whereas the average over many independent draws for $W_{t_{\mathrm{int}}}$ converges on the survival probability, *etc.*.

To calculate the probability of fixation, we are assuming a successful invasion *i.e.* we have conditioned on the non-extinction of the branching process, and evaluate $W_{t_{\mathrm{int}}}$ using the conditional (exponential) distribution. To simplify our computations, we replaced the random initial condition $\frac{W_{t_{\mathrm{int}}}}{n}$ with its mean $\frac{1}{\left(1-Q_{\mathrm{inv}}(t_{\mathrm{int}})\right)n} = \frac{1}{P_{\mathrm{inv}}(t_{\mathrm{int}})n}$ after observing that the latter approximation did not qualitatively change our predictions.

Finally, we note that the proof that, in the limit, this branching process approximation gives the correct invasion probability (*i.e.*, the probability of the number of infectious individuals eventually exceeds $\varepsilon n$) is *mutatis mutandis* that of [8, Proposition 5, Supplementary Information §8.2], : the essential step is to replace bounds on the principal matrix solution linearized at the endemic equilibrium, $\|e^{(t-s)A}\|$, with their counterparts with a time-varying perturbation (see *e.g.*, [11, Theorem 3.20]).

### 5 Invasion and Fixation with Standing Variation

We can compute the probability that some of the vaccine-escape mutations are present as standing variation before the start of vaccination. When the resident population has reached its endemic equilibrium $(X_{\mathrm{u}}(0), 0, Y_{\mathrm{uw}}(0), 0, N(0))$ (SI.1), the number of mutants is approximated by a birth-and-death process with immigration, with birth rate $b_{\mathrm{m}}(0) = \beta_{\mathrm{m}} \frac{X_{\mathrm{u}}(0)}{N(0)}$ (*cf.* (10), but at the wild-type endemic equilibrium $X_{\mathrm{u}}(t) \equiv X_{\mathrm{u}}(0)$ and $X_{\mathrm{v}}(t) \equiv 0$), and death rate $d_{\mathrm{m}} = \delta + \alpha_{\mathrm{m}} + \gamma_{\mathrm{m}}$ (9), and the "immigration" is actually mutations arising in the resident population, which occur at rate $\mu_{\mathrm{m}} = \theta_{\mathrm{u}} Y_{\mathrm{uw}}(0)$. Because we assume that in a fully naïve host population vaccine-escape mutations carry a fitness cost relative to the resident strain, we have $b_{\mathrm{m}}(0) < d_{\mathrm{m}}$. The number of mutants thus approximately follows a subcritical birth-and-death process with immigration, which is known to converge in distribution as $t \to \infty$ to a negative binomial stationary distribution [4]. The probability that there are $j$ infected individuals hosting the mutant pathogen at time $t = 0$ is thus:

$$p_j = \binom{j+k-1}{j}(1-\mathcal{R}_{\mathrm{m}}^{\mathrm{e}}(0))^k(\mathcal{R}_{\mathrm{m}}^{\mathrm{e}}(0))^j \tag{SI.22}$$

where

$$k = \frac{\mu_{\mathrm{m}}}{b_{\mathrm{m}}(0)} \tag{SI.23}$$

is the success (dispersion) parameter and

$$\mathcal{R}_{\mathrm{m}}^{\mathrm{e}}(0) = \frac{b_{\mathrm{m}}(0)}{d_{\mathrm{m}}} = \frac{\beta_{\mathrm{m}} X_{\mathrm{u}}(0)}{\delta + \alpha_{\mathrm{m}} + \gamma_{\mathrm{m}}}. \tag{SI.24}$$

(*cf.* (7)). Hence the the expected number of vaccine-escape mutants already present at the start of vaccination is

$$\frac{\mu_{\mathrm{m}}}{d_{\mathrm{m}} - b_{\mathrm{m}}(0)}. \tag{SI.25}$$

This result is analogous to the classical result that the expected frequency of deleterious mutations is of the form $\mu/s$ where $\mu$ is the rate of mutation towards deleterious mutants and $s$ is the fitness cost of those deleterious mutants.

Using (SI.22), the probability that no mutant is present at the start of vaccination:

$$p_0 = (1-\mathcal{R}_{\mathrm{m}}^{\mathrm{e}}(0))^k \tag{SI.26}$$

When either $\mathcal{R}_{\mathrm{m}}^{\mathrm{e}}(0)$ or $k$ is small then $p_0 \approx 1$ and we can neglect the presence of preexisting mutants.

To compute he probability of adaptation from standing variation at time $t = 0$, we use the probability $p_j$ that there are $j$ mutants present at time $t = 0$ and the estimates for the probabilities of invasion and fixation of a single mutant arriving at time arriving at time $t = 0$ $P_{\mathrm{inv}}(0)$ and $P_{\mathrm{fix}}(0)$, (*cf.* (14) and (33), taking $t_{\mathrm{int}} = 0$). Under the branching process approximation, the chain of infections started by each mutation will go extinct independently with probability $1 - P_{\text{inv}}(0)$. The probability of invasion is then the probability that at least one line survives, $1-(1-P_{\text{inv}}(0))^j$. Summing this over all possible values of $j$ gives us the invasion probability from standing variation,

$$P_{\text{st}} = \sum_{j=1}^{\infty} p_j \left(1 - (1 - P_{\text{inv}}(0))^j\right) = 1 - p_0 - \sum_{j=1}^{\infty} p_j (1 - P_{\text{inv}}(0)))^j = 1 - \sum_{j=0}^{\infty} p_j (1 - P_{\text{inv}}(0)))^j.$$

Recalling that the probability generating function for the number of mutants at time $t = 0$ is

$$\sum_{j=0}^{\infty} p_j z^j = \left(\frac{1 - \mathcal{R}_{\text{m}}^{\text{e}}(0)}{1 - \mathcal{R}_{\text{m}}^{\text{e}}(0) z}\right)^k,$$

which converges provided $|z| < \frac{1}{\mathcal{R}_{\text{m}}^{\text{e}}(0)}$, we see that the probability of invasion from standing variation is $1 - \left(\frac{1-\mathcal{R}_{\text{m}}^{\text{e}}(0)}{1-\mathcal{R}_{\text{m}}^{\text{e}}(0)(1-P_{\text{st}})}\right)^k$.

#### 5.1 Auxiliary Functions for Standing Variation

Similarly, if there are $j$ mutations at time $t = 0$, (22) gives us that $\mathbb{P}\{I_{\text{m}}^n(t) > 0 | Y_{\text{m}}(0) = j\} \approx 1 - (1 - U_{\text{m}}(t|0))^j$, so proceeding as above, we find that the probability that the mutant is still present at time $t$, assuming that at least one individual was present at time $t = 0$ is approximately

$$1 - \left(\frac{1 - \mathcal{R}_{\text{m}}^{\text{e}}(0)}{1 - \mathcal{R}_{\text{m}}^{\text{e}}(0)(1 - U_{\text{m}}(t|0))}\right)^k.$$

Substituting this for $\mathbb{P}\{I_{\text{m}}^n(t) > 0\}$ in (18) and differentiating as above gives us an auxiliary equation analogous to (32) for $U_{\text{st}}(t)$, the probability that the mutant fixes starting from standing variation:

$$\begin{aligned}\dot{U}_{\text{st}} &= \mathbb{P}\{I_{\text{m}}^n(s) > 0\} \frac{d}{dt} \mathbb{P}\{I_{\text{w}}^n(s) = 0\} \\ &= nY_{\text{w}}(0)(\delta + \alpha_{\text{w}} + \gamma_{\text{w}}) U_{\text{st}}^{\text{w}} V_{\text{st}}^{\text{w}} \left(1 - U_{\text{st}}^{\text{w}}\right)^{nY_{\text{w}}(0)-1} \left(1 - \left(\frac{1 - \mathcal{R}_{\text{m}}^{\text{e}}(0)}{1 - \mathcal{R}_{\text{m}}^{\text{e}}(0)(1 - U_{\text{st}}^{\text{m}})}\right)^k\right).\end{aligned}$$

As previously, we obtain

$$P_{\text{fix}}(0) = \lim_{t \to \infty} U_{\text{st}}(t)$$

by choosing $t$ sufficiently large that $|U_{\mathrm{st}}(t+\Delta t) - U_{\mathrm{st}}(t)|$ is smaller than some predetermined error threshold.

#### 5.2 Additional stochastic simulations

We developed a modified version of our simulation approach to relax two simplifying assumptions used throughout our analysis: (i) vaccination starts when the epidemiology has reached an endemic equilibrium, (ii) naturally immune host are fully protected against reinfection. The hybrid approach that we developed could be extended to account for these modifications. But these extensions may obscure the presentation of our work and we prefer to focus here on an exploration of these alternative scenarios with stochastic simulations.

• **The timing of the start of vaccination:** We used a modified version of our simulation code to explore the robustness of our results when vaccination starts sooner and the wild-type pathogen has not reached its endemic equilibrium. Starting vaccination when the incidence has not yet reached an endemic equilibrium tends to increase the probability of invasion of the escape mutant because it is less likely to go extinct when the density of susceptible hosts is more abundant (**Figure SI.5**). Note, however, that this effect is maximised when the mutant is introduced soon after the start of vaccination (*i.e.,* low values of $T_{\mathrm{int}}$). Indeed, for the parameter values we used, the endemic equilibrium is reached very fast and the predictions of the probability of invasion computed when vaccination starts at the endemic equilibrium remain relatively good.

• **Efficacy of natural immunity against reinfections:** We used a modified version of our simulation code to explore the robustness of our results when naturally immune hosts are not perfectly protected. In this scenario, we assume that recovered hosts exposed to strain $i$ have a probability $\epsilon_{\mathrm{r}}\epsilon_i$ to be reinfected. It is still possible to aggregate the density of different types of hosts infected by the same strain $i$, $I_i := I_{\mathrm{u}i} + I_{\mathrm{u}i} + I_{\mathrm{r}i}$, where $I_{\mathrm{r}i}$ refers to the density of recovered hosts rinfected by strain $i$. The effective per-generation reproduction ratio becomes:

$$\mathcal{R}_i^{\mathrm{e}} = \mathcal{R}_i \left( \frac{X_{\mathrm{u}}}{N} + \epsilon_i \frac{X_{\mathrm{v}}}{N} + \epsilon_{\mathrm{r}}\epsilon_i \frac{Z}{N} \right)$$

Hence, higher values of $\epsilon_\mathrm{r}$ give an extra advantage to the escape mutant because it increases the fraction of imperfectly immune hosts. This new expression of the effective per-generation reproduction ratio can be used to derive a good approximation of the probability of mutant invasion using (16). This approximation captures how higher values of $\epsilon_\mathrm{r}$ increase the probability of invasion of escape mutations (**Figure SI.6**). Interestingly, when $\epsilon_\mathrm{r}$ is large, the probability of invasion is less sensitive to the time at which the escape mutant is introduced because, whatever the time after the start of vaccination, the fraction of imperfectly immune host is large.

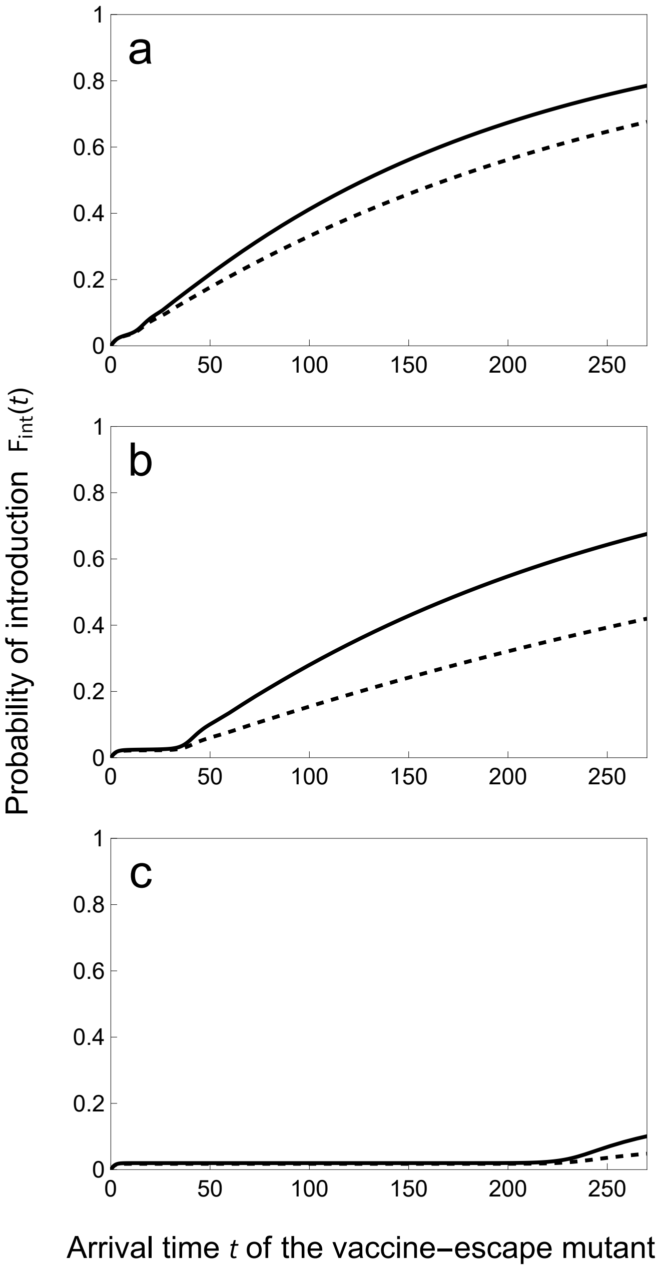


**Figure SI.1: Faster vaccine rollout reduces the probability of introduction of the first escape mutant**. We plot the probability $F_{int}(t)$ that the first escape mutant arrives before time $t$ for different speeds of vaccination rollout: $\upsilon = 0.05$ (top), 0.15 (middle) and 0.24 (bottom). We contrast a scenario where $\theta_{\mathrm{v}} = \theta_{\mathrm{u}}$ (dashed line), and $\theta_{\mathrm{v}} = 10 \times \theta_{\mathrm{u}}$ (full line). Same parameter values as in **Figure 1** in the main text except $\omega_{\mathrm{v}} = 0.05$ and $\omega_{\mathrm{r}} = 0.03$. Note how the lower rate of waning immunity of naturally immune hosts prolongs the drop in the incidence following the start of vaccination (i.e. the “honey moon” period) which yields lower probability of introduction of the escape mutant during this period.

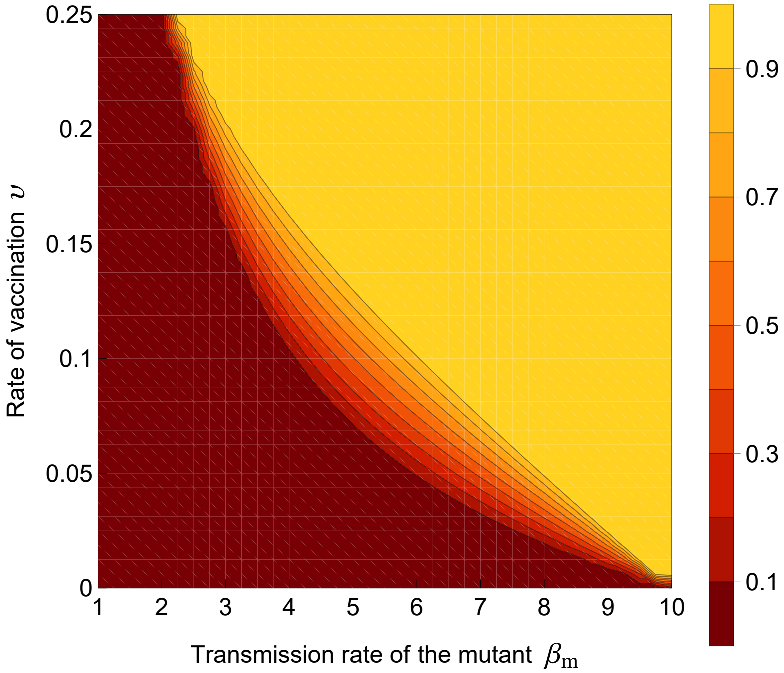


**Figure SI.2: Coexistence between the wildtype and the vaccine-escape mutant for intermediate rates of vaccination**. We plot the equilibrium frequency of the mutant for different rates of transmission $\beta_{\mathrm{m}}$ and for different speeds of vaccination rollout $\upsilon$. Intermediate values of $\beta_{\mathrm{m}}$ and $\upsilon$ promote the coexistence between the two genotypes. Other parameter values: $\nu = \delta = 3\ 10^{-4}$, $\omega_{\mathrm{v}} = \omega_{\mathrm{r}} = 0.05$, $\alpha_{\mathrm{w}} = \alpha_{\mathrm{m}} = 0.02$, $\beta_{\mathrm{w}} = 10$, $\gamma_{\mathrm{w}} = \gamma_{\mathrm{m}} = 2$, $\epsilon_{\mathrm{w}} = 0.05$, $\epsilon_{\mathrm{m}} = 0.05$.

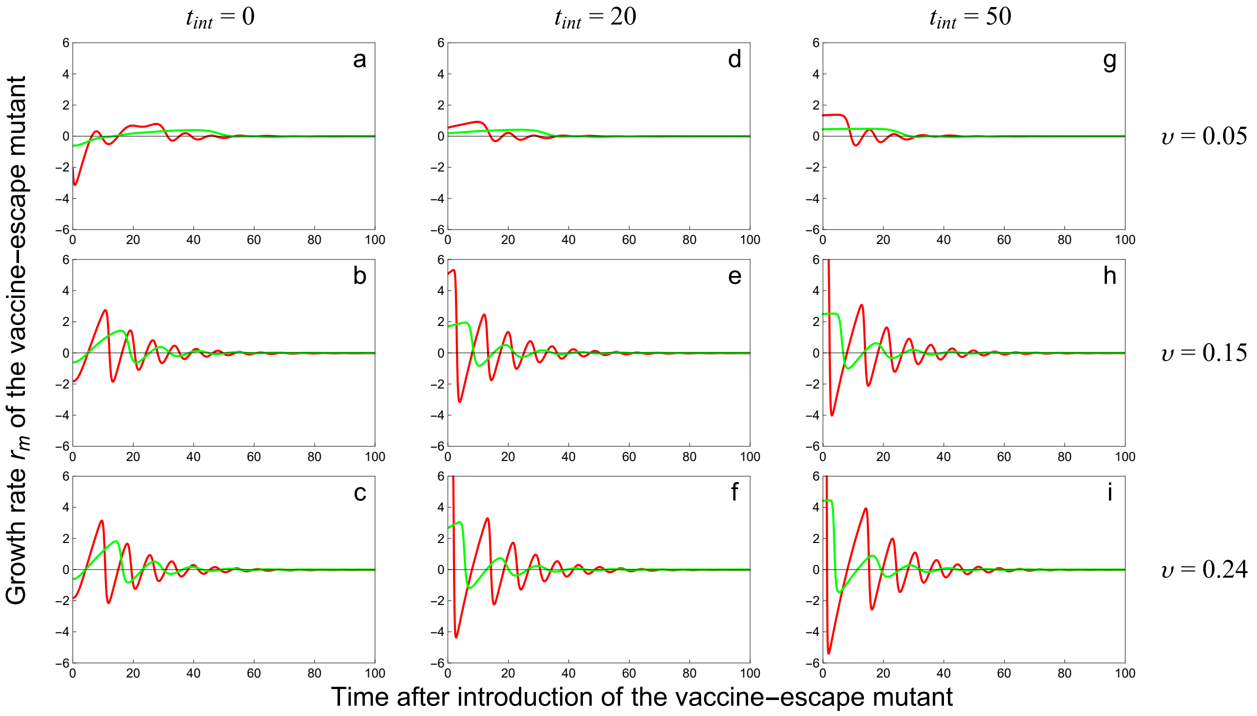


**Figure SI.3: Fluctuations of the per-capita growth rate of the vaccine-escape mutant $\mathcal{R}_{\mathrm{m}}$ for different introduction times $t_{\mathrm{int}}$.** We plot the per-capita growth rate of the vaccine-escape mutant $\mathcal{R}_{\mathrm{m}}$ of a *slow* (green) and a *fast* (red) vaccine-escape mutant for different speeds of vaccination rollout: $\upsilon = 0.05$ (top), 0.15 (middle) and 0.24 (bottom). We also vary the introduction time $t_{\mathrm{int}}$: $t_{\mathrm{int}} = 0$ (left column), 20 (middle column) and 50 (right column). The *slow* mutant: $\alpha_{\mathrm{m}} = 0.02, \beta_{\mathrm{m}} = 7, \gamma_{\mathrm{m}} = 2, \epsilon_{\mathrm{m}} = 1, \mathcal{R}_{\mathrm{m}} = 3.46$. The *fast* mutant: $\alpha_{\mathrm{m}} = 4.0606, \beta_{\mathrm{m}} = 21, \gamma_{\mathrm{m}} = 2, \epsilon_{\mathrm{m}} = 1, \mathcal{R}_{\mathrm{m}} = 3.46$. Other parameter values as in **Figure 3**: $\nu = \delta = 3\ 10^{-4}$, $\omega_{\mathrm{v}} = \omega_{\mathrm{r}} = 0.05$, $\alpha_{\mathrm{w}} = 0.02$, $\beta_{\mathrm{w}} = 10$, $\gamma_{\mathrm{w}} = 2$, $\epsilon_{\mathrm{w}} = 0.05$, $\mathcal{R}_{\mathrm{w}} = 4.95$.

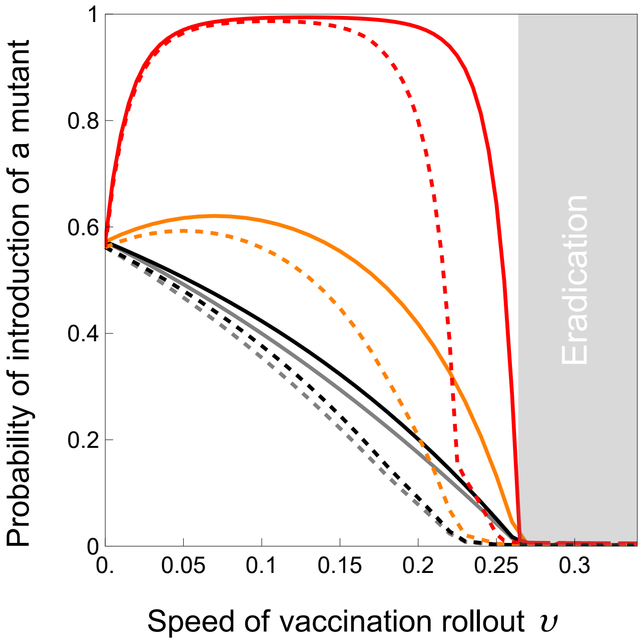


**Figure SI.4: The speed of vaccination rollout affects the probability of introduction of a vaccine-adapted variant by mutation (with or without NPI)**. We plot the probability $F_{int}(t)$ that at least one vaccine-escape variant is introduced by mutation before $t = 1000$. We assume $\theta_{\mathrm{u}} = 0.1$ and we consider four different values of mutation $\theta_{\mathrm{v}}$: $\theta_{\mathrm{v}} = 0.1 \times \theta_{\mathrm{u}}$ (gray), $\theta_{\mathrm{v}} = \theta_{\mathrm{u}}$ (black) $\theta_{\mathrm{v}} = 10 \times \theta_{\mathrm{u}}$ (orange), $\theta_{\mathrm{v}} = 100 \times \theta_{\mathrm{u}}$ (red). The gray area indicates the parameter region where vaccination leads to pathogen eradication (i.e., $\upsilon > \upsilon_{\mathrm{c}} \approx 0.265$, see equation (8) in the main text). The dashed lines correspond to a scenario where we impose periodic fluctuations in $c(t)$, which measures the intensity of Non-Pharmaceutical Interventions (NPIs) that reduce the transmission rate of all pathogens by a fraction $1 - c(t)$. Here we use a square wave function for $c(t)$ that fluctuates between 0.2 and 0 with a period $T = 200$. Other parameter values: $\nu = \delta = 3\ 10^{-4}$, $\omega_{\mathrm{v}} = \omega_{\mathrm{r}} = 0.05$, $\alpha_{\mathrm{w}} = 0.02$, $\beta_{\mathrm{w}} = 10$, $\gamma_{\mathrm{w}} = 2$, $\epsilon_{\mathrm{w}} = 0.05$, $\mathcal{R}_{\mathrm{w}} = 4.95$.

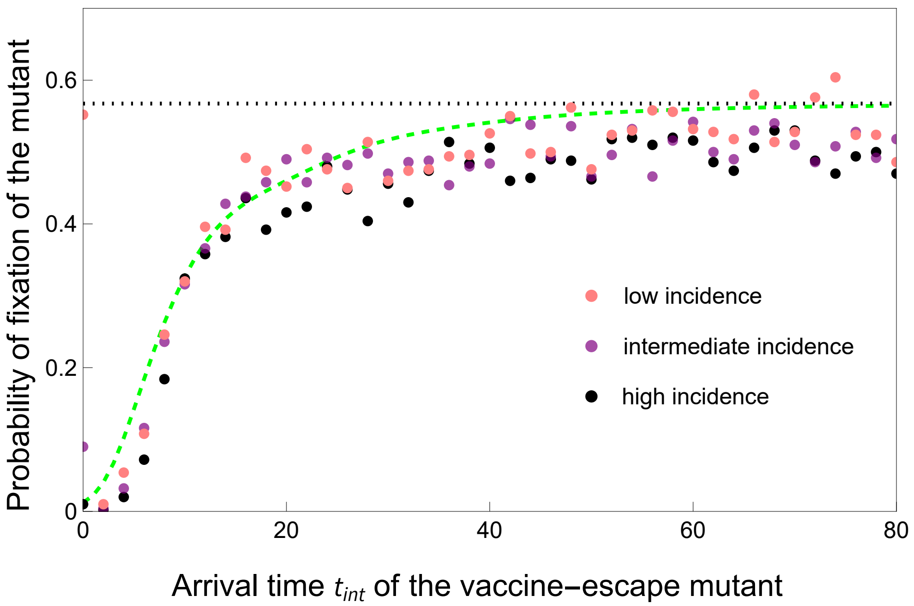


**Figure SI.5: Effect of the incidence of the infection at the start of the vaccination campaign.** We plot the probability invasion $F_{\mathrm{m}}(t_{\mathrm{int}})$ of a *slow* vaccine-escape mutant when the speed of vaccination rollout is $\upsilon = 0.15$ (green dashed line). The dots indicate the value of the probability of fixation obtained from individual based simulation (see §SI- 5.2) for different values of the incidence of the infection at $t = 0$ (i.e. at the start of the vaccination campaign): at low incidence $I^n(0) = 10$ (pink), at intermediate incidence $I^n(0) = 100$ (purple), at high incidence $I^n(0) = 1000$ (black). Note that, for the parameter values we use the endemic incidence is 849 which is close to the high incidence treatment. The *slow* mutant: $\alpha_{\mathrm{m}} = 0.02, \beta_{\mathrm{m}} = 7, \gamma_{\mathrm{m}} = 2, \epsilon_{\mathrm{m}} = 1, \mathcal{R}_{\mathrm{m}} = 3.46$. The probability of invasion $P^{\star}_{\mathrm{m}}$ in the limit $t_{\mathrm{int}} \to \infty$ (see equation (16) in the main text)) is indicated with the dashed black line. Other parameter values: $\nu = \delta = 3\ 10^{-4}$, $n = 10^5$, $\omega_{\mathrm{v}} = \omega_{\mathrm{r}} = 0.05$, $\alpha_{\mathrm{w}} = 0.02$, $\beta_{\mathrm{w}} = 10$, $\gamma_{\mathrm{w}} = 2$, $\epsilon_{\mathrm{w}} = 0.05$, $\mathcal{R}_{\mathrm{w}} = 4.95$.

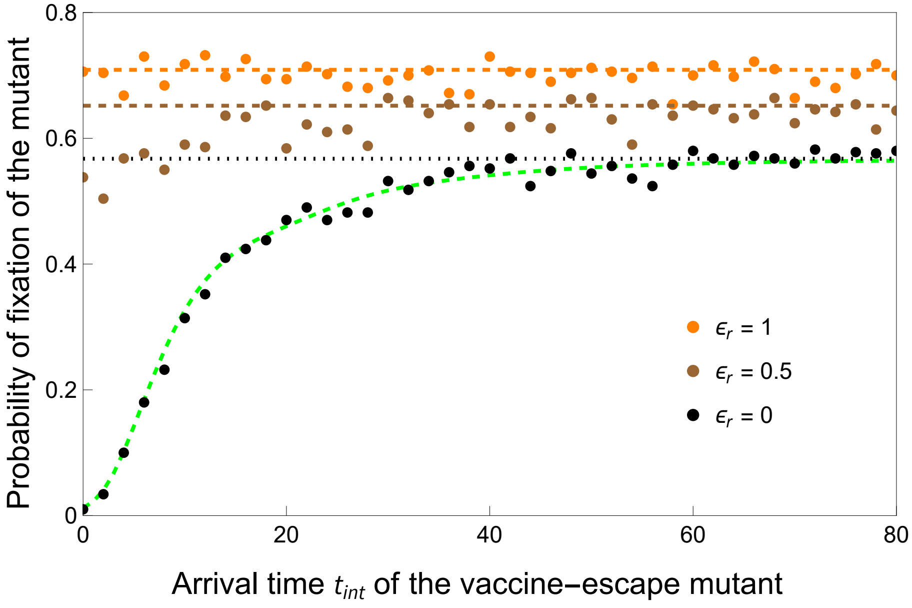


**Figure SI.6: The ability to reinfect naturally immune hosts favors the invasion of the vaccine-escape mutant**. We plot the probability invasion $F_{int}(t_{\rm int})$ of a *slow* vaccine-escape mutant when the speed of vaccination rollout is $\upsilon = 0.15$ and $\epsilon_{\rm r} = 0$ (green dashed line). The *slow* mutant: $\alpha_{\rm m} = 0.02, \beta_{\rm m} = 7, \gamma_{\rm m} = 2, \epsilon_{\rm m} = 1, \mathcal{R}_{\rm m} = 3.46$. The probability of invasion $P_{\rm inv}$ in the limit $t_{\rm int} \to \infty$ (see equation (15) in the main text) is indicated with the dashed black line ($\epsilon_{\rm r} = 0$), the dashed brown line ($\epsilon_{\rm r} = 0.5$) and the dashed orange line ($\epsilon_{\rm r} = 1$). The dots indicate the results of individual based simulation (see §SI-5.2) for different values of $\epsilon_{\rm r} = 0, 0.5, 1$. Other parameter values: $\nu = \delta = 3\ 10^{-4}$, $n = 10^5$, $\omega_{\rm v} = \omega_{\rm r} = 0.05$, $\alpha_{\rm w} = 0.02$, $\beta_{\rm w} = 10$, $\gamma_{\rm w} = 2$, $\epsilon_{\rm w} = 0.05$, $\mathcal{R}_{\rm w} = 4.95$.

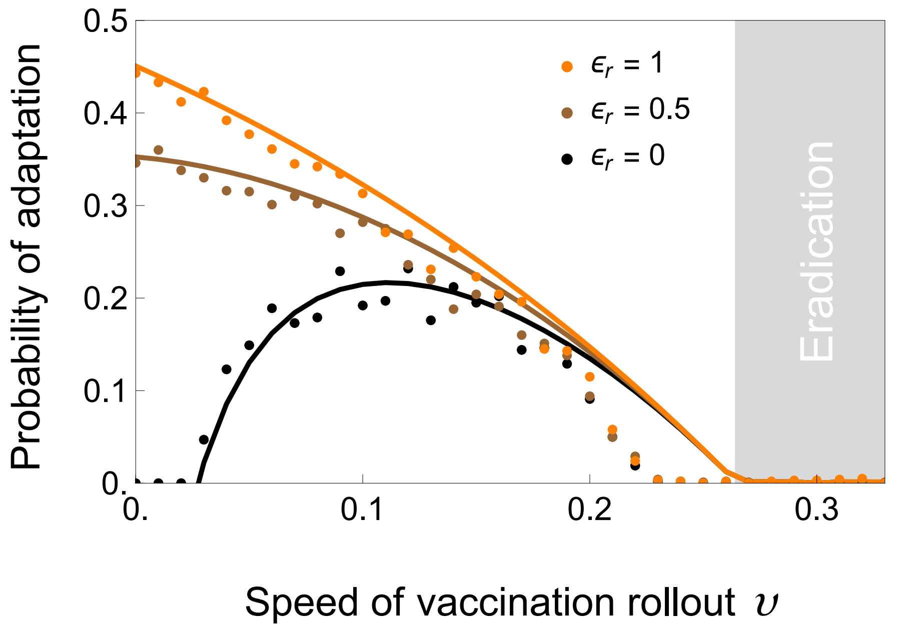


**Figure SI.7: The ability to reinfect naturally immune hosts speeds up the rate of adaptation for low vaccination rollout**. We plot the probability of adaptation $F_{\text{inv}}(t)$ against the speed of vaccination rollout at time $t = 1000$ for different values of $\epsilon_{\text{r}}$: 0 (black), 0.5 (brown), 1 (orange). The dots give the results obtained from individual-based simulations (see §SI- 5.2). The vaccine-escape mutant is assumed to have the following phenotype (*slow* mutant in **Figure 4** and **5**): $\alpha_{\text{m}} = 0.02, \beta_{\text{m}} = 7, \gamma_{\text{m}} = 2, \epsilon_{\text{m}} = 1, \mathcal{R}_{\text{m}} = 3.46$. Other parameter values as in **Figure 6A**: $\nu = \delta = 3\ 10^{-4}$, $n = 10^5$, $\omega_{\text{v}} = \omega_{\text{r}} = 0.05$, $\alpha_{\text{w}} = 0.02$, $\beta_{\text{w}} = 10$, $\gamma_{\text{w}} = 2$, $\epsilon_{\text{w}} = 0.05$, $\mathcal{R}_{\text{w}} = 4.95$. The light gray area on the right-hand-side indicates the speed above which the wild-type pathogen is expected to be driven to extinction ($\upsilon > \upsilon_{\text{c}} \approx 0.264$, see equation (8) in the main text).

**Table SI.1:** Parameters and dynamical variables of the model

| Parameters | | |
|---|---|---|
| Symbol | Description | Default value |
| $\nu$ | influx rate of susceptible hosts | $3\ 10^{-4}$ week$^{-1}$ |
| $\delta$ | natural death rate of hosts | $3\ 10^{-4}$ week$^{-1}$ |
| $\omega_{\mathrm{r}}$ | rate of waning immunity of naturally immune hosts | 0.05 week$^{-1}$ |
| $\omega_{\mathrm{v}}$ | rate of waning immunity of vaccinated hosts | 0.05 week$^{-1}$ |
| $\upsilon$ | rate of vaccination | variable |
| $\theta_{\mathrm{u}}$ | rate of mutation (from w to m) in unvaccinated hosts | variable |
| $\theta_{\mathrm{v}}$ | rate viral mutation (from w to m) in vaccinated hosts | variable |
| $n$ | system size (scaling parameter allowing us to manipulate the pathogen population size) | variable |
| $\alpha_{\mathrm{w}}$ | virulence (increased mortality rate) by strain w | 0.02 week$^{-1}$ |
| $\beta_{\mathrm{w}}$ | rate of transmission of strain w | 10 week$^{-1}$ |
| $\gamma_{\mathrm{w}}$ | recovery rate of the host infected by strain w | 2 week$^{-1}$ |
| $\epsilon_{\mathrm{w}}$ | probability of infection of vaccinated hosts by strain w | 0.05 |
| $\mathcal{R}_{\mathrm{w}}$ | reproduction number of strain w | 4.95 |
| $\alpha_{\mathrm{m}}$ | virulence (increased mortality rate) by strain m | *variable* |
| $\beta_{\mathrm{m}}$ | rate of transmission of viral strain m | *variable* |
| $\gamma_{\mathrm{m}}$ | recovery rate of the host infected by strain m | 2 week$^{-1}$ |
| $\epsilon_{\mathrm{m}}$ | probability of infection of vaccinated hosts by strain m | 1 |

| Variables | | |
|---|---|---|
| Number | Description | Density |
| $S_{\mathrm{u}}^n$ | unvaccinated susceptible hosts | $X_{\mathrm{u}}$ |
| $S_{\mathrm{v}}^n$ | vaccinated susceptible hosts | $X_{\mathrm{v}}$ |
| $R^n$ | recovered hosts | $Z$ |
| $I_{\mathrm{u}i}^n$ | unvaccinated hosts infected by strain $i \in \{\mathrm{w}, \mathrm{m}\}$ | $Y_{\mathrm{u}i}$ |
| $I_{\mathrm{v}i}^n$ | vaccinated hosts infected by strain $i \in \{\mathrm{w}, \mathrm{m}\}$ | $Y_{\mathrm{v}i}$ |
